# From simulation to pedagogy: structured AI standardized patients for clinical communication training validated through multi-model and randomized evaluation

**DOI:** 10.64898/2026.04.26.26351793

**Authors:** Ping Wu, Yu Han, Jing Zhang, Yunqi Li, Mengna Jiang, Xinyu Lu, Haibin Zhang, Danyang Xu, Hao Ming, Lihong Wang, Qingping Wen

## Abstract

Standardized patients (SPs) are central to clinical communication training but are constrained by cost, scalability, and reliance on trained actors. We present AI standardized patients (AI-SPs), large language model–driven simulators governed by a three-layer information architecture that modulates disclosure according to learner skill. We validate this approach across three phases. In Phase 1, blinded expert evaluation of 350 simulated consultations from five frontier LLMs showed that learner skill level, rather than model choice, drove performance variation (***η*^2^ = 0.31** vs **0.06**), indicating that pedagogical quality emerges from architectural design rather than model scaling. In Phase 1b, 155 live student consultations revealed systematic failures in eliciting safety-critical information, generating automated curriculum diagnostics without expert observation. In a three-arm pilot randomized controlled trial (Phase 2, ***n* = 58**), AI-SP training achieved skill gains non-inferior to human SP practice, with a distinctive self-efficacy benefit unique to the AI-SP arm. These findings suggest that architecture-driven AI-SPs offer a scalable, model-portable paradigm for clinical communication training.

## 1 Introduction

Clinical communication skills are fundamental to patient safety, yet their development requires deliberate practice with realistic patient encounters[1, 2]. Standardized patients (SPs) remain the gold standard[3, 4], but their dependence on trained actors, scheduling coordination, and faculty supervision creates resource barriers that constrain training frequency, particularly in specialties such as anaesthesiology, where preoperative consultations demand integration of history-taking, risk communication, and emotional support within high-stakes, time-pressured encounters[5, 6].

Large language models (LLMs) offer a potential solution. Current models demonstrate clinical knowledge[7, 8], conversational fluency[9], and the capacity to adopt complex personas[10], capabilities that surpass prior virtual patient systems built on branching scripts or constrained natural language processing[11, 12]. Early work has shown promise for generative AI in personalised medical education[13] and clinical communication[14, 15]. Recent studies have advanced AI-based patient simulation: Luo et al.[16] demonstrated LLM-based history-taking training in ophthalmology; Schwill et al.[17] reported a two-arm RCT showing AI-simulated clinical encounters improved OSCE performance over standard training; and Yu et al.[18] developed an LLM-based simulated patient agent validated with medical students. However, these studies share important limitations: all used a single LLM platform (raising generalisability concerns), none compared AI-SP directly against human SP practice (the clinical gold standard), and none employed a structured pedagogical framework governing information disclosure. Creating educationally effective AI-SPs requires more than conversational fluency; it demands pedagogically controlled information disclosure that rewards skilled interviewing, emotional responses calibrated to communication quality, and clinical consistency across conversational turns[19].

Here we introduce a *three-layer information architecture* for AI-SP design that addresses this gap. Surface information (Layer 1) is volunteered spontaneously; prompted information (Layer 2) is disclosed only upon direct questioning; and hidden information (Layer 3) is withheld, not through deliberate concealment but because the simulated patient lacks the health literacy to recognise its clinical relevance, requiring targeted, empathetic questioning to elicit (Fig. 1A1). This design mirrors documented patient behaviour in which critical clinical details emerge only through skilled communication[20], and operationalises cognitive apprenticeship theory[21] by generating a calibrated gradient between what novice and competent students can discover. Crucially, the hidden-information discovery rate provides an objective, automated metric of clinical communication skill without requiring real-time expert observation, enabling scalable assessment alongside scalable practice. The iceberg metaphor (Supplementary Fig. 7) further visualises this graduated disclosure: Layer 1 sits above the waterline accessible to all students, while Layer 3 resides in deep water, discoverable only through empathetic probing that builds sufficient patient trust.

**Fig. 1.**
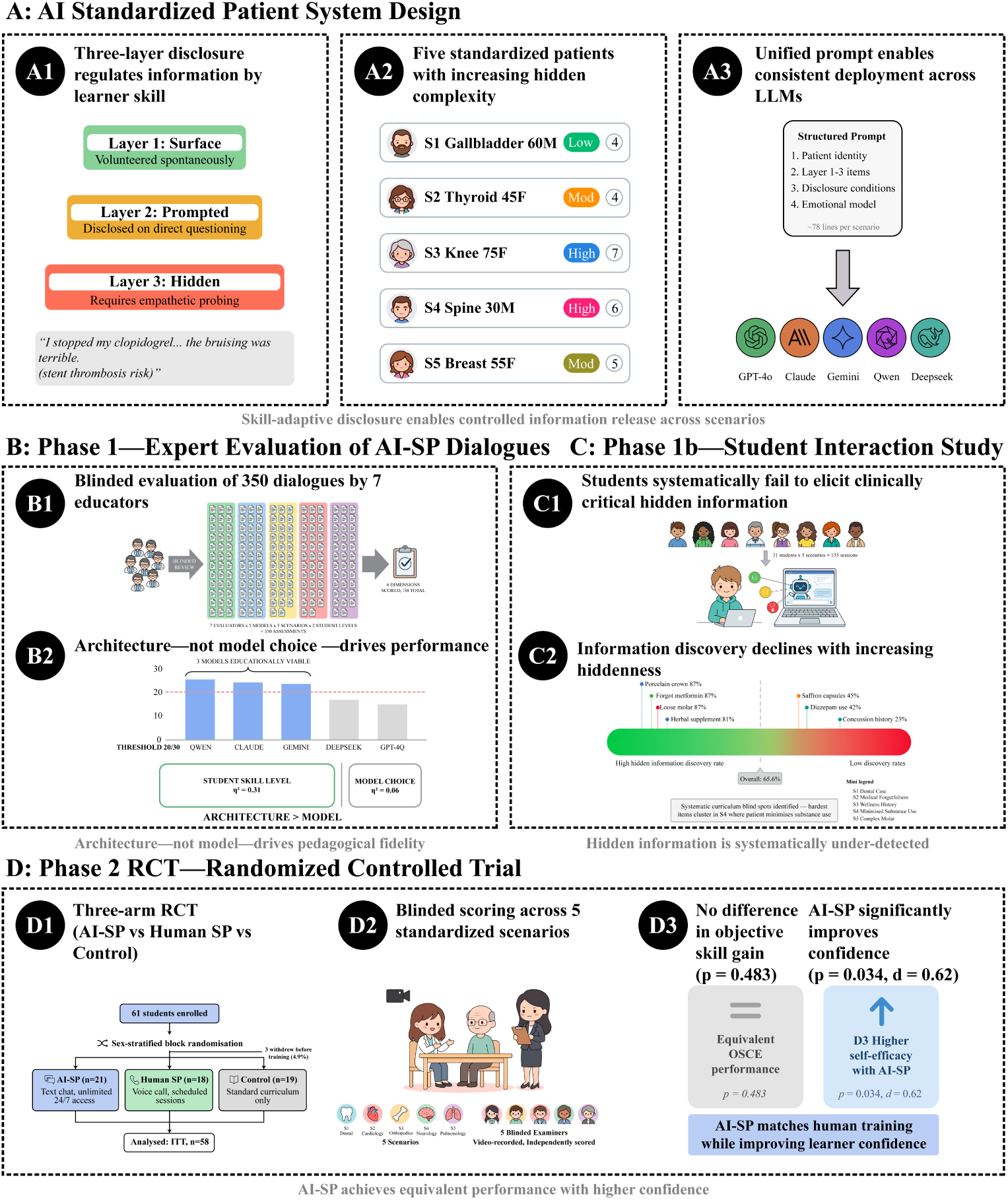
AI standardized patient system design and three-phase validation overview. **(A)** System design: (A1) Three-layer information architecture: Layer 1 (surface) is volunteered spontaneously, Layer 2 (prompted) requires direct questioning, Layer 3 (hidden) requires empathetic probing to unlock safety-critical information; (A2) five preoperative scenarios spanning low to high complexity with safety-critical hidden items; (A3) unified prompt enabling consistent deployment across LLMs. **(B)** Phase 1, expert evaluation of AI-SP dialogues: (B1) blinded evaluation of 350 dialogues by seven educators; (B2) architecture, rather than model choice, drives performance. **(C)** Phase 1b, student interaction study: (C1) students systematically fail to elicit clinically critical hidden information; (C2) information discovery declines with increasing hiddenness. **(D)** Phase 2, randomized controlled trial: (D1) three-arm RCT (AI-SP vs human SP vs control); (D2) blinded scoring across five stan-dardized scenarios; (D3) no difference in objective skill gain (*p* = 0.48) while AI-SP significantly improves self-efficacy confidence (*p* = 0.034, *d* = 0.62).

We evaluate this approach through a three-phase progressive validation pipeline targeting preoperative anaesthesia consultations (Fig. 1B–D). This pipeline follows a deliberate escalation logic: deploying an AI training system in a clinical education setting without first establishing its fidelity and ecological validity would be methodologically premature and ethically questionable. Each phase therefore addresses a prerequisite question that must be answered before the next phase can be justified:

- **Phase 1 (Construct validity):** *Can AI-SPs produce educationally adequate sim-ulations?* Seven anaesthesiology educators blindly evaluated dialogues from five frontier LLMs (GPT-4o, Claude 4.5 Sonnet, Gemini 2.5 Flash, Qwen-2.5 Plus, DeepSeek-R1) across five clinical scenarios at two student skill levels, yielding 350 assessments.
- **Phase 1b (Ecological validity):** *Do scripted-dialogue findings generalise to authentic student interactions?* Thirty-one students completed 155 real-time con-sultations to characterise interaction quality and per-item discovery patterns under uncontrolled conditions.
- **Phase 2 (Training efficacy):** *Does AI-SP practice improve clinical skills com-parably to human SP practice?* A three-arm pilot RCT (*n* = 58) compared AI-SP, human SP, and standard curriculum controls on OSCE performance and self-efficacy.

Translating the three-layer architecture into a system that functions reliably across multiple LLMs required a structured prompt engineering process (Fig. 2). Over four iterative development rounds with expert review, a clinician–engineer team identified and resolved recurring failure modes, including role boundary violations, premature information disclosure, affective flattening, and stylistic artefacts, producing a final six-section prompt structure that is identical across all five LLMs tested.

**Fig. 2.**
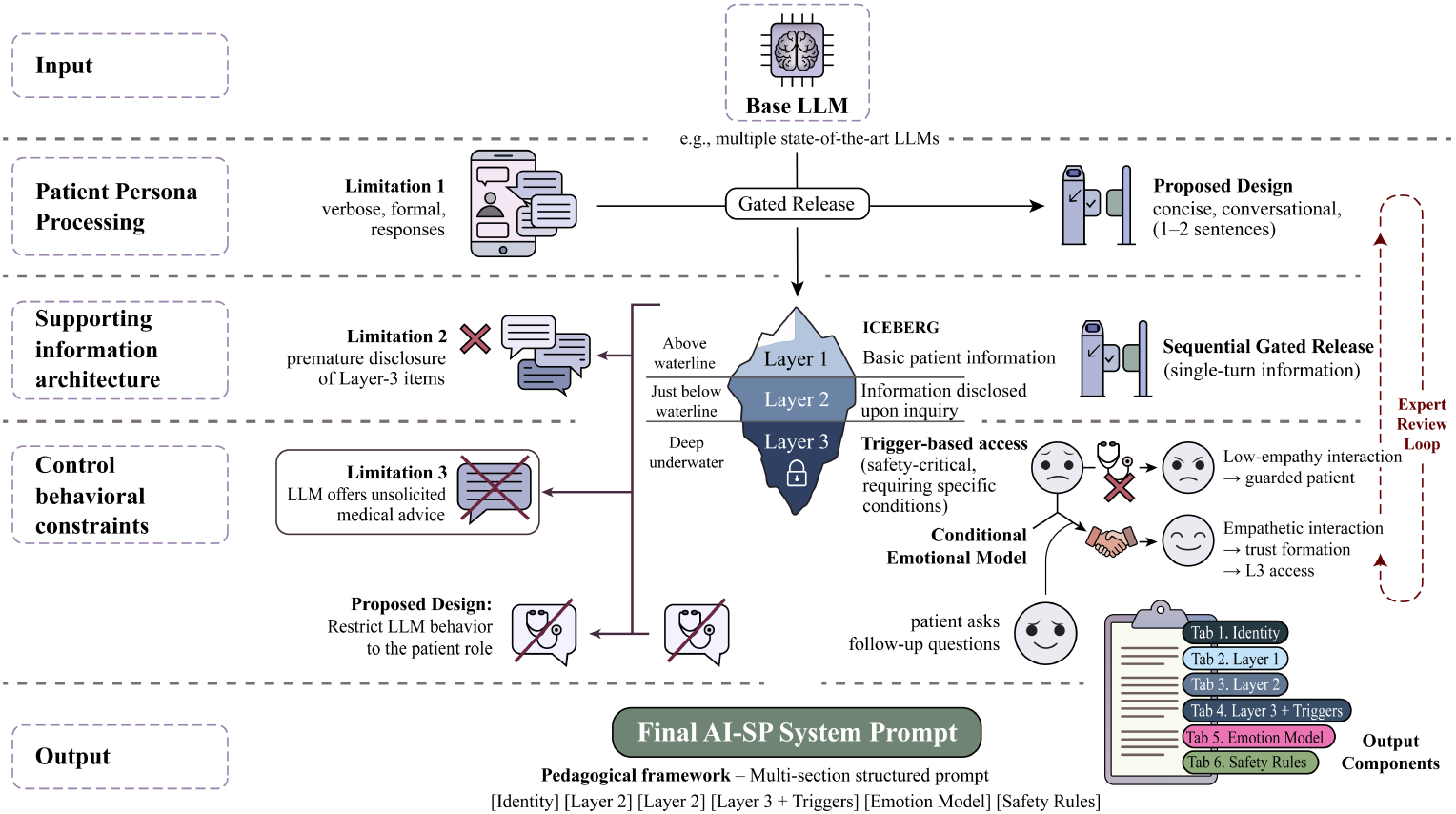
AI standardized patient prompt engineering framework. Starting from a base LLM, four iterative development rounds addressed distinct failure modes: verbose and formal patient responses (persona calibration), premature disclosure of safety-critical Layer 3 information (gated release with explicit trigger conditions), flat emotional affect regardless of interviewer quality (conditional emotional model linking empathy to trust and information access), and role boundary violations where the LLM offered unsolicited medical advice (behavioural constraints restricting the model to the patient role). Expert review after each round refined the prompt. The final system prompt comprises six structured sections (identity, Layers 1–3 with trigger conditions, emotional model, and safety rules), averaging 78 lines per scenario, deployed identically across all five LLMs.

## 2 Results

Results are presented following the progressive validation sequence: first establishing that AI-SPs can produce educationally adequate simulations (Phase 1), then confirming these findings generalise to authentic student interactions (Phase 1b), and finally testing whether AI-SP practice translates to measurable training outcomes (Phase 2).

### 2.1 Phase 1: Expert validation of AI-SP fidelity

#### Question: Can AI-SPs produce educationally viable simulations, and does fidelity depend on the LLM or the pedagogical architecture?

Blinded expert evaluation revealed significant between-model differences (*p <* 0.001), with a two-tier structure (Fig. 3a; Table 1): three models (Qwen-2.5 Plus, Claude 4.5 Sonnet, Gemini 2.5 Flash) exceeded the pre-specified educational viability threshold, while DeepSeek-R1 and GPT-4o fell below it. Post-hoc comparisons confirmed that the top-tier models significantly outperformed the lower tier (Bonferroni-corrected *p <* 0.025), while within-tier differences were non-significant.

**Fig. 3.**
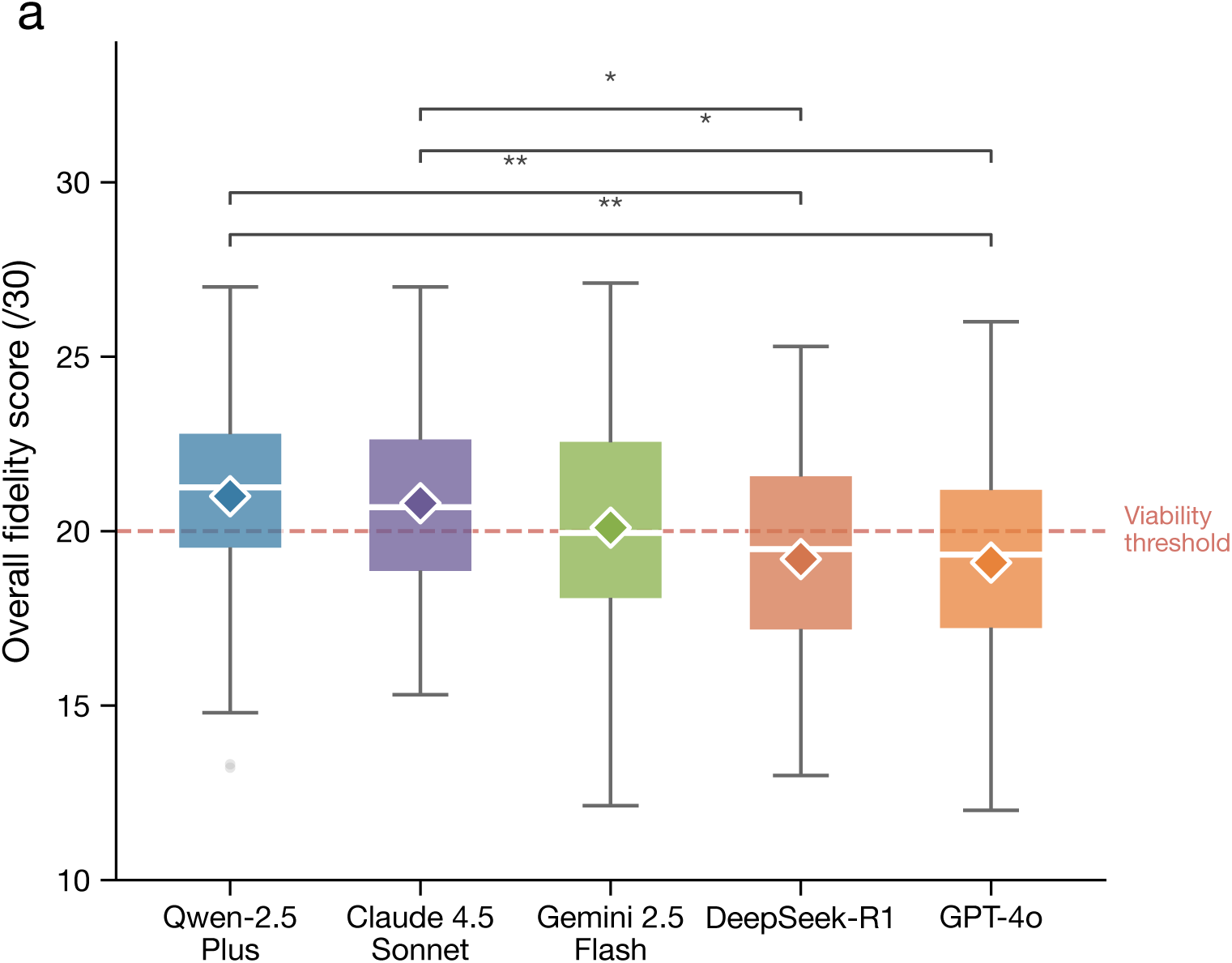
Phase 1 expert fidelity evaluation (*n* = 7 evaluators, 350 assessments). **a**, Overall fidelity by model. Box plots show median and IQR; diamonds indicate means; dashed line marks the educational viability threshold (≥20/30). Significance brackets: Bonferroni-corrected (^∗^*p <* 0.05; ^∗∗^*p <* 0.01). **b**, Fidelity comparison between competent and novice student interactions across six dimensions. Error bars: SEM. **c**, Fidelity heatmap across models and scenario complexity levels (Low: S1; Mod-erate: S2, S5; High: S3, S4). Green dots indicate cells ≥20/30 (educationally viable).

**Table 1.** AI-SP fidelity scores across six evaluation dimensions (mean ± SD, scale 1–5). Total scores sum to 30.

| Model | Clin.<br>real. | Emot.<br>auth. | Info.<br>ctrl. | Comm.<br>nat. | Behav.<br>cons. | Pedag.<br>val. | Total<br>(/30) |
| --- | --- | --- | --- | --- | --- | --- | --- |
| Qwen-2.5 Plus | **3.51±0.70** | **3.60±0.75** | **3.34±0.59** | **3.59±0.71** | 3.60±0.62 | 3.39±0.71 | **21.0±2.9** |
| Claude 4.5 Sonnet | 3.53±0.65 | 3.41±0.73 | 3.26±0.56 | 3.46±0.77 | **3.63±0.57** | **3.53±0.61** | 20.8±2.9 |
| Gemini 2.5 Flash | 3.53±0.76 | 3.33±0.77 | 3.20±0.67 | 3.41±0.79 | 3.46±0.72 | 3.19±0.77 | 20.1±3.5 |
| DeepSeek-R1 | 3.23±0.75 | 3.21±0.66 | 3.23±0.59 | 3.20±0.73 | 3.30±0.64 | 3.07±0.79 | 19.2±3.1 |
| GPT-4o | 3.10±0.85 | 3.19±0.75 | 3.21±0.61 | 3.13±0.70 | 3.33±0.61 | 3.13±0.80 | 19.1±3.3 |

Bold indicates highest score per dimension. Educational viability threshold: $\geq 20/30$ ( $\geq 3.33$ mean per dimension). Three models exceeded this threshold; DeepSeek-R1 and GPT-4o fell slightly below.

The key finding was that student skill level, rather than model choice, drove fidelity variation. Two-way ANOVA confirmed that student level accounted for five-fold more variance than model identity (*η*^2^ = 0.31 vs 0.06), with no significant interaction (*p <* 0.001; Table 2; Fig. 3b). The largest differentiation occurred in educational value and communication naturalness.

**Table 2.**
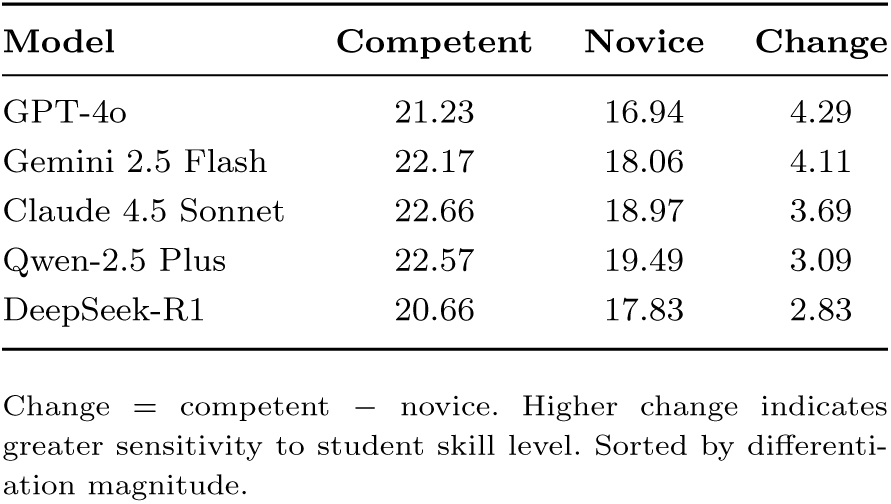
AI-SP fidelity scores by student interaction level (total score /30).

| Model | Competent | Novice | Change |
| --- | --- | --- | --- |
| GPT-4o | 21.23 | 16.94 | 4.29 |
| Gemini 2.5 Flash | 22.17 | 18.06 | 4.11 |
| Claude 4.5 Sonnet | 22.66 | 18.97 | 3.69 |
| Qwen-2.5 Plus | 22.57 | 19.49 | 3.09 |
| DeepSeek-R1 | 20.66 | 17.83 | 2.83 |

Change = competent – novice. Higher change indicates greater sensitivity to student skill level. Sorted by differentiation magnitude.

This gradient extended to hidden information disclosure (Table 3): competent students discovered all 26 hidden Layer 3 items, while novice students discovered only three, none of which were safety-critical. Missed items included self-discontinued clopidogrel creating stent thrombosis risk (S3), unreported ephedrine-containing supplements (S4), and saffron capsules posing serotonin syndrome risk with concurrent sertraline (S5).

**Table 3.** Hidden information (Layer 3) discovery rates by scenario and student level.

| Scenario | Hidden items | *n* | Competent | Novice |
| --- | --- | --- | --- | --- |
| S1 (Low) | Aspirin, alcohol, snoring, loose molar | 4 | 4/4 (100%) | 0/4 (0%) |
| S2 (Mod.) | Herbal thyroid tea, dental prosthesis, missed metformin, unresolved cold | 4 | 4/4 (100%) | 0/4 (0%) |
| S3 (High) | Self-stopped clopidogrel, palpitations, husband death, OSA, memory decline, dentures, penicillin allergy | 7 | 7/7 (100%) | 1/7 (14.3%) |
| S4 (High) | Daily ibuprofen, fitness supplements (ephedrine), concussion, heavy drinking, benzodiazepine use, e-cigarettes | 6 | 6/6 (100%) | 0/6 (0%) |
| S5 (Mod.) | Saffron capsules (5-HT risk), double-dose lorazepam, extreme diet, advance directive, smoking | 5 | 5/5 (100%) | 2/5 (40.0%) |
| **Total** |  | **26** | **26/26 (100%)** | **3/26 (11.5%)** |

Novice students failed to discover any safety-critical hidden items across all five scenarios. *n* = number of hidden Layer 3 items per scenario.

Fidelity was robust across scenario complexity (Fig. 3c). Top-performing models maintained scores near the viability threshold even for high-complexity scenarios, with decrements of *<*1 point from low to high complexity. GPT-4o showed the largest complexity-related decline (change score = 1.2).

### 2.2 Phase 1b: Ecological validation through live student interactions

#### Question: Do Phase 1 findings hold when real students interact freely with AI-SPs, and can the system generate actionable curriculum diagnostic data?

To assess whether scripted-dialogue findings generalise to authentic interactions, 31 medical students each completed all five scenarios, generating 155 live consultations. Sessions exhibited features absent from scripted dialogues, including spontaneous follow-up questions, unexpected tangents, premature topic transitions, and natural communication errors.

The overall hidden-information discovery rate was 65.6%, with substantial inter-scenario variation (Table 4): S2 (thyroidectomy) achieved the highest rate and S4 (lumbar disc) the lowest, consistent with the difficulty of eliciting information that patients minimise.

**Table 4.** Phase 1b live student–AI-SP interaction results by scenario (*n* = 31 students, 155 sessions).

| Scenario | Hidden items | Discovery rate | Items found | Messages | Duration |
| --- | --- | --- | --- | --- | --- |
| S1 Cholecystectomy | 4 | 65.3% ± 31.4% | 2.6/4 | 45 ± 12 | 15.4 min |
| S2 Thyroidectomy | 4 | 85.5% ± 34.0% | 3.4/4 | 34 ± 9 | 14.9 min |
| S3 Knee replacement | 7 | 67.3% ± 37.8% | 4.7/7 | 38 ± 13 | 14.5 min |
| S4 Lumbar disc | 6 | 51.1% ± 36.2% | 3.1/6 | 37 ± 17 | 10.7 min |
| S5 Mastectomy | 5 | 58.7% ± 42.2% | 2.9/5 | 35 ± 13 | 13.3 min |
| **Overall** | **26** | **65.6% ± 37.8%** | — | **38 ± 14** | **13.8 min** |

Discovery rate = proportion of hidden Layer 3 items disclosed during the interaction. Values are mean ± SD across 31 students.

Per-item analysis revealed wide variation in discovery rates (Fig. 4). Items related to common clinical concerns (S2) were near-universally discovered, while the hardest items clustered in S4 (where the patient minimised substance use), with concussion history proving the most elusive. Student-level variation was similarly wide, with a clear separation between top-and bottom-quartile performers.

**Fig. 4.**
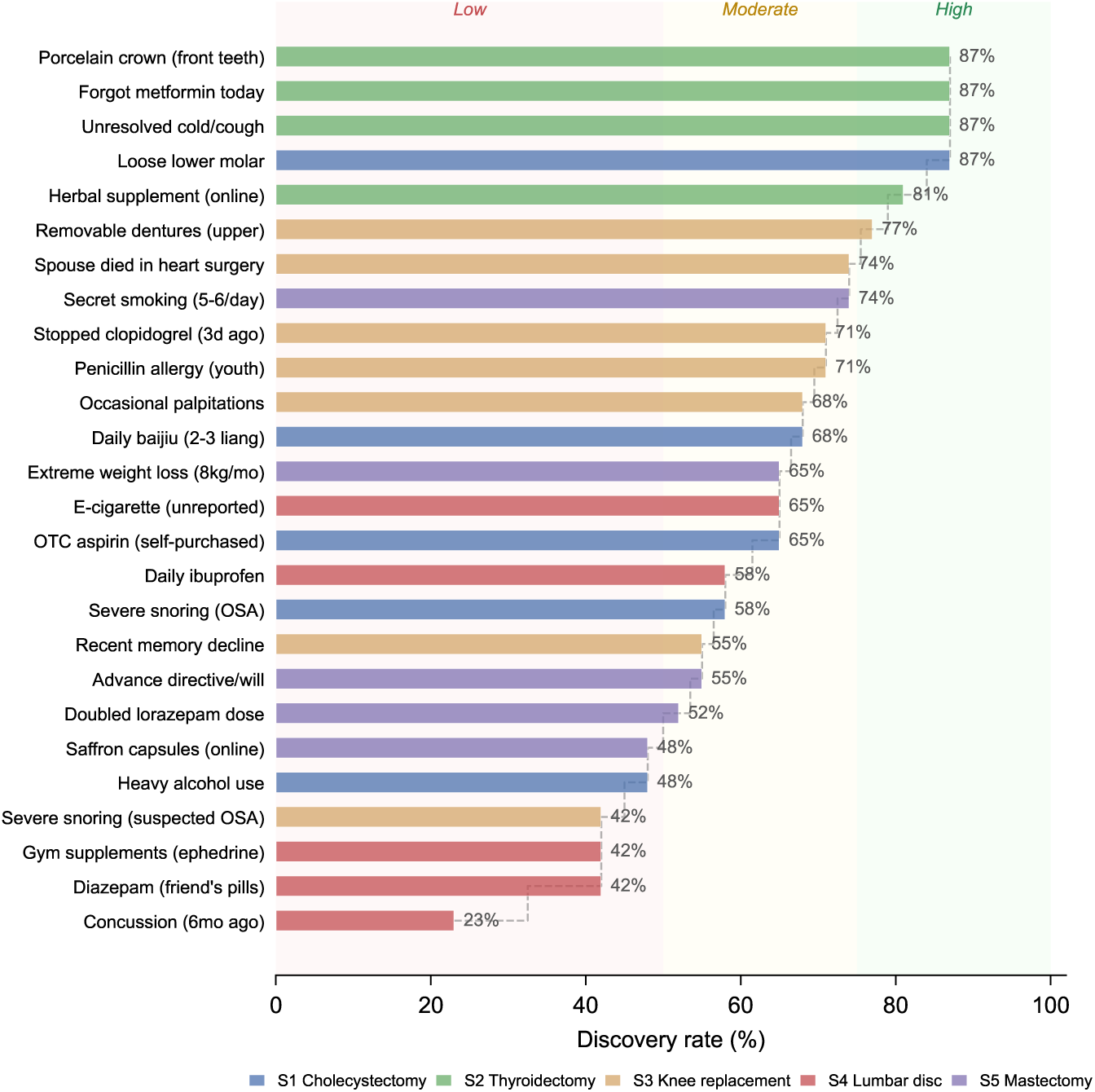
Per-item hidden-information discovery rates across five scenarios (*n* = 31 students, 26 items). Items ranked by discovery rate and colour-coded by scenario. S2 items were near-universally discovered (*>*80%) while S4 items, particularly concussion history (23%) and diazepam use (42%), proved most challenging, reflecting the difficulty of eliciting information that patients actively minimise.

A post-training questionnaire indicated high overall satisfaction (Table 5; Cronbach’s *α* = 0.85). Students rated learning effectiveness and communication skill improvement most highly, while their understanding of the three-layer architecture received the lowest score, suggesting that the pedagogical mechanism operated effectively without being transparent to learners.

**Table 5.** Phase 1b post-training questionnaire results (1–5 Likert scale, *n* = 21 respondents).

| Item | Mean ± SD |
| --- | --- |
| Learning effectiveness | 4.76 ± 0.44 |
| Communication skill improvement | 4.71 ± 0.46 |
| Overall satisfaction | 4.52 ± 0.60 |
| Comparable to human SP | 4.33 ± 0.73 |
| Understanding of three-layer architecture | 3.67 ± 0.97 |

### 2.3 Phase 2: Randomized controlled trial

#### Question: Does AI-SP training produce skill gains equivalent to human SP practice, and does it confer any unique benefits?

Sixty-one students were randomized via sex-stratified block randomization; three withdrew before training, yielding 58 analysed participants across three arms (Fig. 5). Groups were comparable at baseline on all measures (Table 6).

**Fig. 5.**
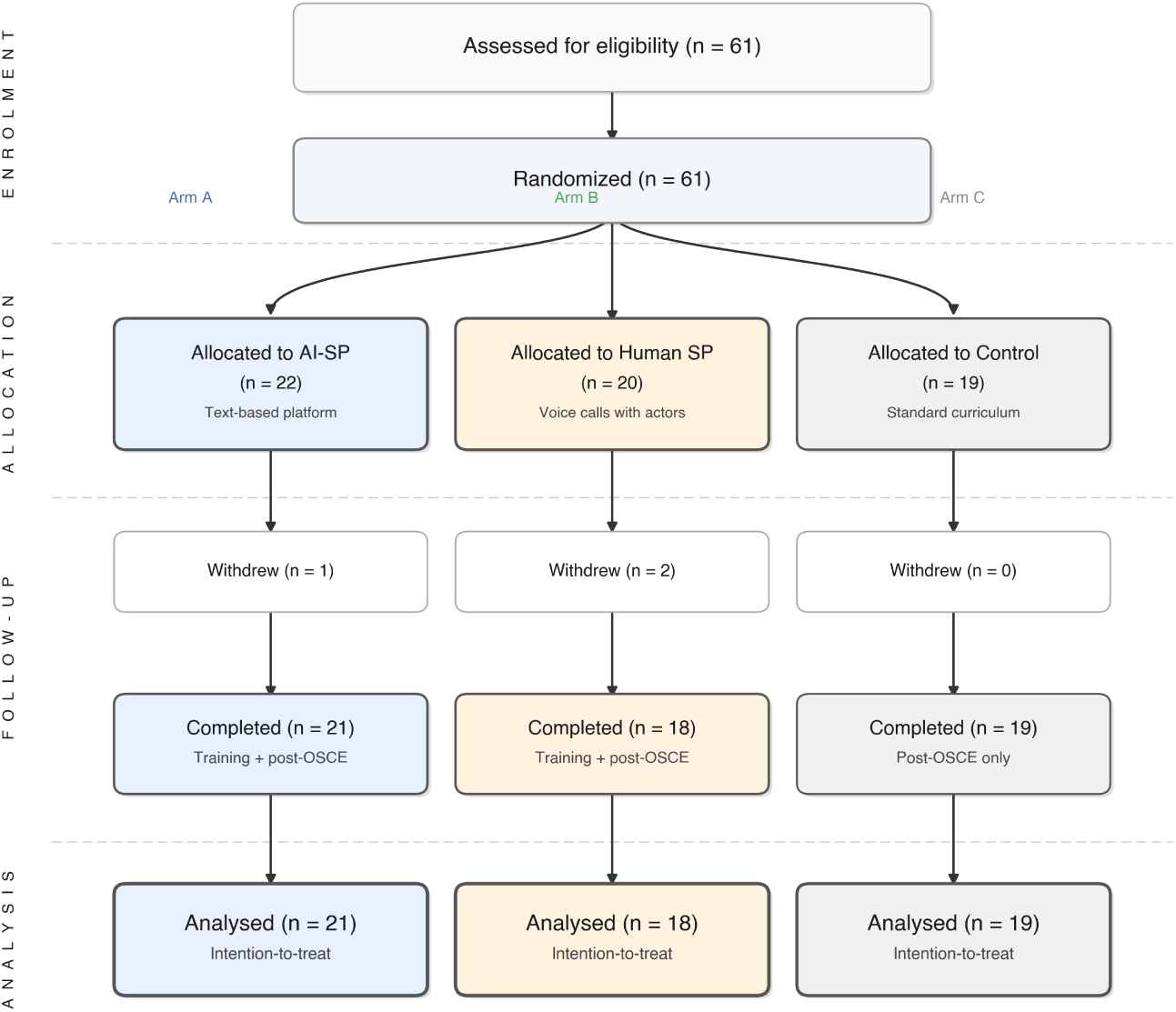
CONSORT flow diagram for the Phase 2 pilot RCT. Sixty-one students were randomized into three arms; three withdrew before training commenced (attrition rate: 4.9%). All 58 remaining participants completed the study protocol and were included in intention-to-treat analysis.

**Table 6.** Phase 2 RCT outcomes: baseline characteristics and pre–post changes across study arms.

| Outcome | AI-SP<br>(*n* = 21) | Human SP<br>(*n* = 18) | Control<br>(*n* = 19) | *p* | Effect size |
| --- | --- | --- | --- | --- | --- |
| *Baseline (pre-training)* |  |  |  |  |  |
| OSCE checklist (/14) | 7.5±1.1 | 7.7±1.3 | 7.2±1.5 | 0.715 | — |
| OSCE total (/39) | 18.5±1.8 | 19.1±2.7 | 18.3±2.6 | 0.588 | — |
| Self-efficacy (/50) | 25.1±9.1 | 30.6±10.1 | 27.3±9.5 | 0.193 | — |
| *Primary outcome* |  |  |  |  |  |
| Checklist change (ICC=0.72) | +1.4±1.0 | +0.9±1.3 | +1.0±1.2 | 0.483<sup>a</sup> | *d*<sub>AB</sub>=0.38 |
| *Secondary outcomes* |  |  |  |  |  |
| OSCE total change (ICC=0.40) | +8.6±1.9 | +7.3±3.4 | +7.6±2.6 | 0.266<sup>a</sup> | *d*<sub>AB</sub>=0.52 |
| Self-efficacy change | +10.2±6.3 | +6.8±6.8 | +6.1±6.8 | 0.064<sup>a</sup> | *d*<sub>AC</sub>=0.62* |
| Satisfaction (/55)<sup>†</sup> | 45.0±7.2 | 44.4±6.9 | — | 0.876<sup>b</sup> | *d*<sub>AB</sub>=0.09 |

Values are mean ± SD. <sup>a</sup>Kruskal-Wallis H test across three arms. <sup>b</sup>Mann-Whitney U test (two arms). <sup>†</sup>Satisfaction collected for intervention arms only (AI-SP: *n* = 21; Human SP: *n* = 16; two Human SP participants did not return the satisfaction questionnaire). \* *p* = 0.034, AI-SP vs Control pairwise comparison. *d*<sub>AB</sub>: Cohen's *d* for AI-SP vs Human SP; *d*<sub>AC</sub>: AI-SP vs Control.

Inter-rater reliability differed markedly by scoring instrument: the binary checklist achieved good reliability (ICC = 0.72) while the global rating scale showed poor reliability (ICC = 0.15; Fig. 6). Accordingly, the checklist score was designated as the primary outcome, with the total score reported as secondary (full ICC values with 95% CIs in Supplementary Tables 7–8).

**Fig. 6.**
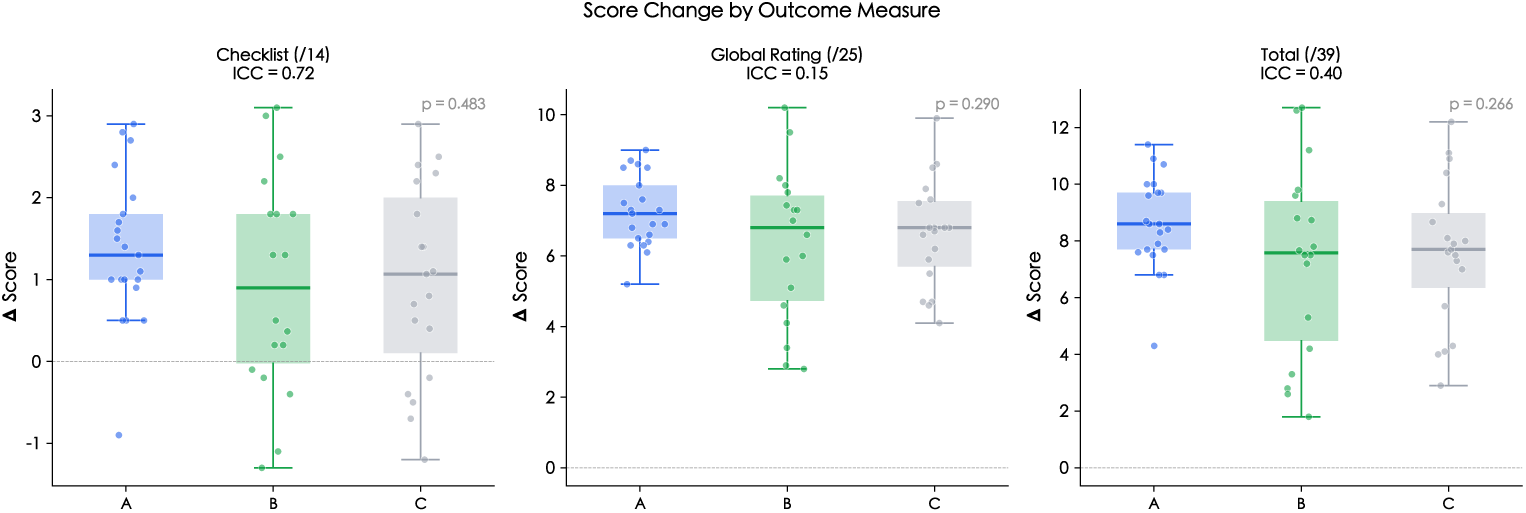
Phase 2 score changes by outcome measure. Each panel shows pre–post score change by study arm (box plots with individual data points; ⋄ = mean). **a**, Checklist score (/14; ICC = 0.72; primary outcome): all groups improved comparably (*p* = 0.483). **b**, Global rating score (/25; ICC = 0.15): poor inter-rater reliability motivated its exclusion as primary outcome. **c**, Total OSCE score (/39; ICC = 0.40): consistent pattern of equivalence across arms (*p* = 0.266).

**Fig. 7.**
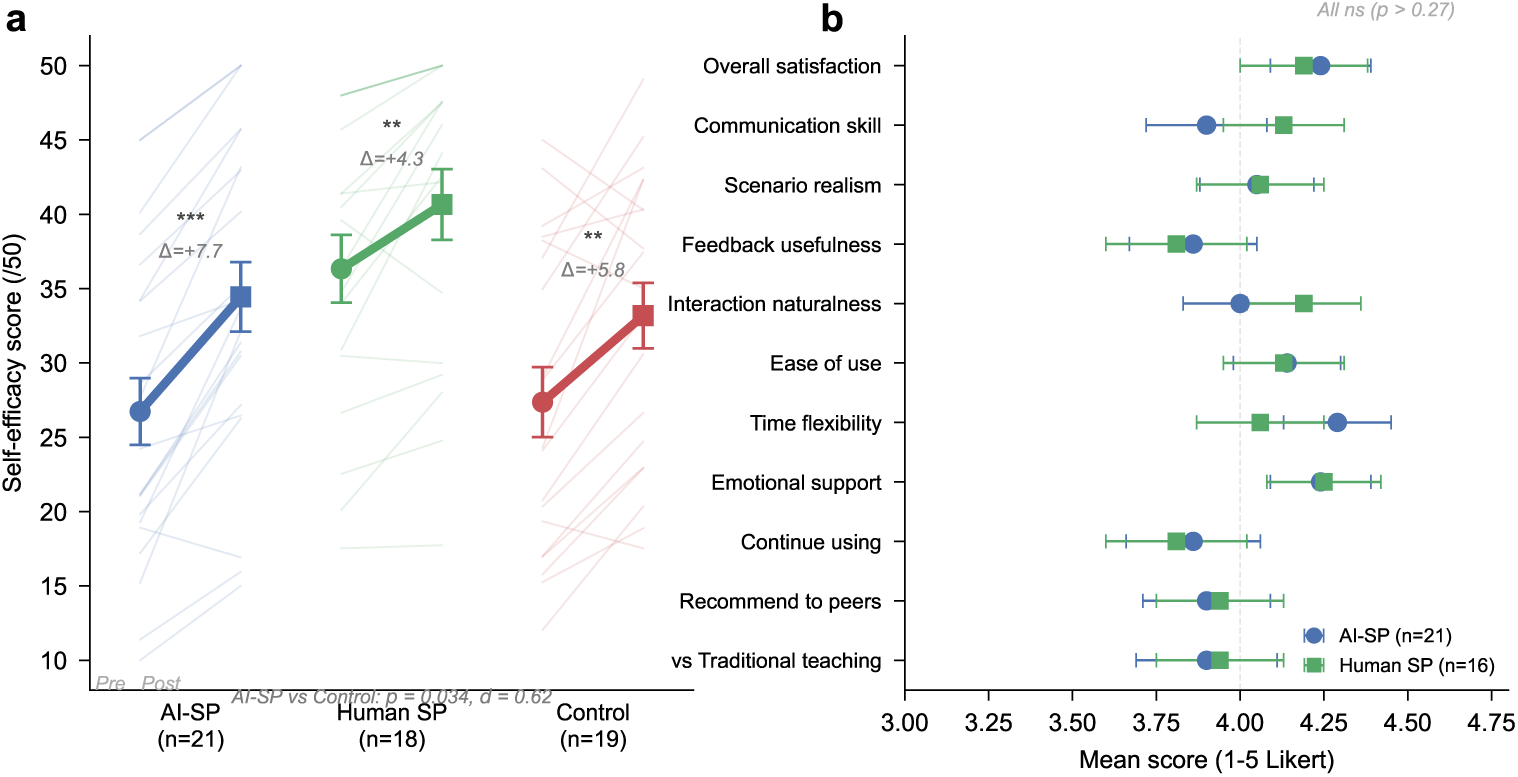
Phase 2 secondary outcomes. **a**, Self-efficacy scores (/50) before and after training. Individual student trajectories (thin lines) and group means (± SEM; circles = pre, squares = post). All groups improved significantly (^∗∗^*p <* 0.01; ^∗∗∗^*p <* 0.001), but the AI-SP group showed the largest mean gain (mean change = +10.2), significantly exceeding controls (*p* = 0.034, *d* = 0.62). **b**, Per-item training satisfaction comparison between AI-SP and human SP arms (1–5 Likert scale). No item showed significant between-group differences (all *p >* 0.27); interaction naturalness numerically favoured human SP, while time flexibility favoured AI-SP.

### Primary outcome: OSCE checklist score

All three groups improved significantly from baseline, with no significant between-group difference (Table 6; Fig. 6a). The AI-SP group showed the numerically largest checklist improvement, but the effect size versus human SP was small. The control group’s comparable improvement suggests a substantial testing effect from repeated OSCE exposure. ANCOVA confirmed equivalence after adjusting for baseline scores, which alone explained nearly half of post-training variance, leaving limited residual variance for training-specific effects.

### Secondary outcome: total OSCE score

The total OSCE score (checklist + global rating) showed a consistent pattern: all groups improved substantially with no significant between-group difference (Fig. 6c; Table 6), paralleling the checklist finding with a medium AI-SP versus human SP effect size.

### Self-efficacy

Self-efficacy improved in all groups, but the AI-SP group showed significantly greater gains than controls, with a trend toward superiority over human SP (Table 6). Individual trajectories (Fig. 7a) reveal a striking pattern: AI-SP students showed nearly uniformly upward shifts, whereas human SP and control groups exhibited greater variability, including several students whose self-efficacy declined. This tighter post-training distribution suggests that unlimited, low-stakes practice benefited weaker students disproportionately. Per-item analysis (Supplementary Table 5; Supplementary Fig. 5) revealed the largest AI-SP advantages in managing time pressure and handling special cases, domains where repeated practice without performance anxiety is most likely to build confidence.

### Satisfaction

Training satisfaction was equivalent between arms (Table 6). Per-item comparison (Fig. 7b) reveals a nuanced pattern beneath the overall equivalence: AI-SP scored numerically higher on time flexibility and emotional support (affordances of asynchronous, judgement-free practice), while human SP scored higher on interaction naturalness, the one dimension where voice-based interaction has an inherent advantage. That the two modalities achieved near-identical satisfaction despite these complementary strengths suggests students valued different aspects of each format. Both arms scored above 4.0/5.0 on all dimensions, with high willingness to continue using AI-SP and recommend it to peers, indicating acceptability for sustained deployment.

## 3 Discussion

This study demonstrates that structured AI standardized patients, governed by a three-layer information architecture, can achieve expert-validated fidelity, produce meaningful diagnostic data about student clinical communication skills, and, when used for training, yield skill outcomes equivalent to both human SP practice and test-retest learning, with a distinctive self-efficacy benefit unique to the AI-SP arm. Three findings merit particular discussion.

### 3.1 Architecture over model: implications for deployment

The most consequential finding is that the variance attributable to student skill level (*η*^2^ = 0.31) was five-fold larger than that attributable to model choice (*η*^2^ = 0.06). This indicates that the three-layer information architecture, rather than the underlying LLM, is the primary driver of pedagogical differentiation. The practical implication is that institutions need not invest in the most expensive commercial API to achieve educationally adequate AI-SP performance; the architecture itself provides the pedagogical mechanism, with the LLM serving as a replaceable engine. Indeed, three models spanning distinct accessibility tiers (a Chinese-developed model, Qwen-2.5 Plus; a premium Western model, Claude 4.5 Sonnet; and a free-tier model, Gemini 2.5 Flash) all exceeded the viability threshold, while the open-source DeepSeek-R1 enables fully self-hosted deployment without API dependency[22]. This landscape means resource-constrained programmes can select from multiple viable options and focus investment on instructional design rather than model selection.

That said, model choice is not irrelevant. In our Chinese-language context, Qwen’s native training data conferred advantages in communication naturalness and emotional authenticity, dimensions requiring culturally calibrated patient behaviour including colloquial expressions and age-appropriate speech patterns. GPT-4o’s lower fidelity despite strong clinical knowledge[23] highlights that benchmark superiority does not guarantee culturally authentic patient simulation, a finding relevant for non-English medical education contexts worldwide.

### 3.2 From assessment tool to curriculum diagnostic

The three-layer architecture’s 88.5 percentage-point discovery differential between competent and novice students validates it as a genuine pedagogical mechanism that operationalises cognitive apprenticeship principles[21]: just as real patients share sensitive information selectively based on interaction quality[20], AI-SPs withhold Layer 3 items until students demonstrate the requisite communication skills. This mechanism offers three advantages over traditional SP training. First, the hidden-information discovery rate provides an *objective, automated assessment metric* without requiring expert observation. Second, AI-SPs apply disclosure rules deterministically, creating a *consistent assessment standard* that human SPs (whose disclosure behaviour may vary despite training[3]) cannot match. Third, *per-item difficulty profiling* generates granular curriculum diagnostic data that would be difficult to obtain at scale with human SPs.

The Phase 1b ecological validation demonstrated this diagnostic capacity: per-item discovery rates ranging from 23% (concussion history) to 87% (porcelain crown) across 26 items revealed that students reliably inquire about conditions they associate with anaesthetic risk but consistently miss information that patients minimise or normalise. The concentration of low-discovery items in S4, where a young male patient normalises recreational supplement use and conceals benzodiazepine dependence, identifies a specific curricular gap in training students to recognise and probe patient minimisation behaviours. These findings extend the paradigm demonstrated by Luo et al.[16] in ophthalmology by showing that AI-SP systems can provide not only training but also systematic diagnostic data about educational blind spots[24].

### 3.3 Equivalent skills, superior confidence: the case for complementary deployment

The Phase 2 RCT reveals a clear pattern: all three groups (AI-SP, human SP, and untrained controls) achieved comparable OSCE skill gains, while the groups diverged sharply on self-efficacy. This combination of skill equivalence with psychological differentiation carries important implications.

The absence of between-group differences in OSCE performance (*p* = 0.483; checklist *d_AB_* = 0.38) constitutes preliminary equivalence evidence: AI-SP practice achieved comparable skill outcomes to the clinical gold standard of human SP training. This is not a null finding but a clinically meaningful result, because AI-SP practice operates at marginal costs orders of magnitude below human SP programmes; if outcomes are equivalent, the cost-effectiveness argument alone justifies deployment. The dominant source of improvement was a testing effect: ANCOVA confirmed that baseline scores alone explained 48.5% of post-training variance, leaving limited variance for training-specific effects to capture[25]. Importantly, the three-arm design allows contextualisation that two-arm studies cannot: the largest published AI-SP RCT (Schwill et al., *n* = 247[17]) reported a significant AI advantage but compared AI only against standard training without a human SP arm, a design that cannot distinguish AI efficacy from the absence of an active comparator. Our pilot RCT (*n* = 58), though smaller, provides a structurally more informative comparison: the inclusion of both human SP and no-intervention arms enables simultaneous superiority (vs control) and equivalence (vs gold standard) inferences. This is a qualitative design advantage, not merely a quantitative limitation. The pilot was designed to estimate effect sizes for a future confirmatory trial[26, 27] rather than to detect small differences; post-hoc power analysis (*N* ≥ 216 needed for 80% power at *d* = 0.38[28]) confirms that definitive equivalence testing requires a larger multicentre study. Critically, the three-phase design means the RCT is contextualised by 350 expert assessments and 155 live student sessions; the total evidence base supporting AI-SP validity is substantially larger than the Phase 2 sample alone.

Where AI-SP training clearly differentiated was in self-efficacy: AI-SP students gained significantly more confidence than controls (mean change = +10.2 vs +6.1; *p* = 0.034, *d* = 0.62), with a trend toward superiority over human SP (mean change = +10.2 vs +6.8; *p* = 0.069). Bandura’s self-efficacy theory[29] identifies four sources of efficacy beliefs: mastery experience, vicarious experience, verbal persuasion, and physiological/affective states. AI-SP training may amplify two of these. First, the psychologically safe environment, where errors carry no interpersonal consequences and practice can be repeated at self-selected times, reduces performance anxiety (affective state), enabling students to accumulate positive mastery experiences without the social evaluation apprehension inherent in human SP encounters. Second, unlimited repetition provides more frequent mastery experiences per unit time than scheduled human SP sessions. The finding that managing time pressure showed the largest per-item AI-SP advantage (*p* = 0.060) supports this interpretation: students who rehearse without time anxiety develop greater confidence in time management specifically. Self-efficacy is not merely a subjective outcome; in clinical education, it predicts willingness to attempt challenging patient interactions, persistence when communication breaks down, and ultimately clinical performance in unsupervised settings[4]. Notably, the modality difference (text vs voice) may also contribute: text-based interaction allows more deliberate response composition, potentially reinforcing a sense of communicative competence that transfers to self-efficacy ratings.

Training satisfaction was equivalent between AI-SP and human SP arms (*p* = 0.876), with both scoring above 4.0/5.0 on all dimensions. This finding contrasts with early concerns that students would resist AI-mediated clinical training, and suggests that the educational value and convenience of AI-SP practice compensate for any perceived limitations in interaction naturalness.

Notably, the entire Phase 2 protocol (training, assessment, and data collection) was conducted fully remotely. AI-SP training used text-based chat; human SP training used voice calls via videoconferencing software with trained actors following identical scripts; OSCE assessments were video-recorded and scored asynchronously by blinded examiners. This demonstrates that an effective AI-SP training programme can be deployed without dedicated simulation facilities, on-site scheduling infrastructure, or synchronous examiner coordination, a deployment pathway particularly relevant for programmes serving geographically distributed learners and infeasible with traditional in-person SP programmes. The practical implication is straightforward: if AI-SP training produces equivalent skills, superior confidence, and equivalent satisfaction at a fraction of the cost, then the barrier to wider adoption is no longer evidence but implementation.

### 3.4 Limitations and future directions

Several limitations warrant consideration. First, the AI-SP and human SP arms used different communication modalities (text-based chat vs voice calls), introducing a confound that prevents attributing outcome differences solely to the AI vs human distinction. This was a deliberate design choice reflecting current deployment reality: text-based LLM interaction is the most mature, accessible, and reproducible modality available today, while human SP training is inherently voice-based; forcing it into text would misrepresent the comparator and reduce ecological validity. Matching modalities would improve internal validity but at the cost of testing a comparison that no institution would actually implement. Notably, text-based interaction is conventionally considered a communicative disadvantage relative to voice, as it lacks paralinguistic cues, real-time turn-taking, and tonal modulation that are central to clinical communication[30, 31]. The finding that text-based AI-SP achieved equivalent OSCE outcomes *despite* this modality disadvantage strengthens rather than weakens the case for AI-SP deployment: if equivalence holds under disadvantaged conditions, it is likely to hold or improve as multimodal AI-SPs incorporating voice and video interaction mature. Future studies should include a text-based human SP arm and a voice-based AI-SP arm to formally disentangle modality and agent effects.

Second, the Phase 2 RCT (*n* = 58) was designed and powered as a pilot study; while baseline equivalence was confirmed, the sample was insufficient to detect small between-group OSCE differences (post-hoc power *<* 25% for observed *d* = 0.38; *N* ≥ 216 needed for 80% power). The absence of a significant OSCE difference therefore reflects both genuine equivalence and limited statistical power, a distinction that only a larger confirmatory trial can resolve.

Third, within-group variability in OSCE score changes was substantial (checklist change score SD = 1.0–1.3 on a 14-point scale), compounded by a strong testing effect that elevated all groups’ scores and compressed between-group differences. We addressed this by: (a) using ANCOVA to adjust for baseline scores, which accounted for 48.5% of post-training variance; (b) pre-specifying the high-reliability checklist (ICC = 0.72) as the primary outcome; and (c) reporting both parametric (ANCOVA) and non-parametric (Kruskal-Wallis, Mann-Whitney) tests. Fourth, using the same five scenarios for both training and assessment introduced scenario-specific familiarity effects. Future implementations should employ a larger scenario bank with non-overlapping training and assessment scenarios to isolate genuine communication skill transfer from scenario memorisation.

Fifth, the study assessed immediate post-training outcomes only; longer-term skill retention (4–12 weeks) and transfer to real patient encounters were not evaluated. Sixth, inter-rater reliability for the global rating scale was poor (ICC = 0.15), necessitating reliance on the checklist score as the primary outcome. While binary checklist items are inherently more reliable than subjective Likert scales, this limits assessment to observable behaviours rather than holistic communication quality. Low global rating reliability is a well-documented challenge in OSCE assessment: Regehr et al.[32] reported global rating ICCs of 0.20–0.40 across clinical tasks, and Gershov et al.[33] recently reported comparable variability in AI-assisted competency assessment. Our decision to report ICC transparently and conduct three layers of robustness analysis (leave-one-out, sensitivity, examiner variability; Supplementary Note 1, Supplementary Fig. 5) represents a methodological strength. Enhanced examiner calibration with video-anchored exemplars should be prioritised in future studies.

Additional methodological considerations include: the study was conducted at a single institution with Chinese-language scenarios, limiting cross-linguistic and cross-cultural generalisability. However, the three-layer information architecture is language-agnostic and domain-agnostic by design (the disclosure rules operate at the level of communication behaviour, not language syntax) and the five-model comparison already embeds deployment diversity across US and Chinese providers, commercial and open-source platforms, and three accessibility tiers. The remote video-conferenced OSCE format, while validated in post-pandemic literature[34, 35], does not assess paralinguistic and non-verbal communication skills. A multicentre, multilingual replication across specialties is planned as the next phase of this research programme.

Taken together, this study addresses three gaps in the emerging AI-SP literature. Unlike prior single-model studies[16, 17, 36], the five-model comparison establishes model-agnostic evidence for the pedagogical architecture. Unlike two-arm designs that compare AI-SP to standard training, our three-arm RCT includes the clinically meaningful human SP comparator alongside a no-intervention control, enabling both superiority (vs control) and equivalence (vs human SP) inferences. And unlike technical benchmarking studies[18], the three-phase validation pipeline progressively establishes construct validity, ecological validity, and training efficacy. Future research should prioritise adequately powered multicentre equivalence trials with extended follow-up, cross-specialty validation, investigation of optimal AI-SP–human-SP integration ratios in curricula, and development of multimodal AI-SPs incorporating voice and video interaction. The finding that AI-SP training uniquely boosts self-efficacy warrants dedicated investigation into the psychological mechanisms underlying this effect. The three-layer information architecture may also be applicable beyond medical education to any professional training context where skilled questioning is central, including journalism, social work, and legal interviewing.

## 4 Methods

### 4.1 Study design and ethics

This three-phase study was conducted at the Department of Anesthesiology, The First Affiliated Hospital of Dalian Medical University, China. Phase 1 validated AI-SP fidelity through blinded expert evaluation of scripted dialogues. Phase 1b extended validation to live student–AI-SP interactions. Phase 2 employed a three-arm, parallel-group randomized controlled trial. The study was reported following DECIDE-AI guidelines[37] for AI system evaluation and CONSORT guidelines for the RCT component. The study was determined to be exempt from formal ethics review by the Ethics Committee of The First Affiliated Hospital of Dalian Medical University (exemption reference: YJ-MS-2025-02; document ID: AF/SW-31/2023-01.0), as it involved comparison of standard educational methods among medical students with no patient contact, no clinical intervention, and no collection of biological specimens. All student participants provided written informed consent.

### 4.2 Scenario design and AI-SP construction

Five preoperative anaesthesia consultation scenarios were developed through iterative expert consensus, stratified by complexity: S1 (low: laparoscopic cholecystectomy, 60M), S2 (moderate: thyroid nodulectomy, 45F), S3 (high: total knee replacement, 75F), S4 (high: lumbar disc surgery, 30M), and S5 (moderate: mastectomy, 55F). Scenarios covered the spectrum of preoperative communication challenges including medication reconciliation, substance use disclosure, psychosocial assessment, and management of acute anxiety.

Each AI-SP was constructed with a patient persona and the three-layer information architecture described in the Introduction (Fig. 1A1). Each scenario contained 4–7 hidden Layer 3 items, including at least one safety-critical item whose non-discovery would represent a clinically significant assessment gap (Table 3). System prompts were structured in four sections: *patient identity* (demographics, personality, speech patterns), *Layer 1 items* (volunteered spontaneously), *Layer 2 items* (disclosed when directly asked), and *Layer 3 items with disclosure conditions* (gated behind specific communication behaviours). Additionally, each prompt specified an emotional response model defining baseline anxiety, triggers for escalation or de-escalation, defensive reactions to insensitive questioning, and gradual trust-building responses to empathetic communication[1, 38]. All five LLMs received identical system prompts. Model parameters were standardised (temperature = 0.7, max tokens = 4096, top p = 0.9). Translating this architecture into effective system prompts required iterative prompt engineering (Fig. 2; Supplementary Table 2). Over four development rounds, a clinician–engineer team tested each draft prompt against all five LLMs across both competent and novice student interaction patterns, identifying and resolving four recurring failure modes: (i) *role boundary violations*, where the model offered differential diagnoses or treatment suggestions rather than remaining in the patient role; (ii) *premature information disclosure*, where multiple Layer 3 items were revealed in a single turn regardless of conversational context; (iii) *affective flattening*, where the model responded with uniformly cooperative behaviour irrespective of the student’s communication quality; and (iv) *stylistic artefacts*, including verbose, formal phrasing inconsistent with the patient persona and bracketed action descriptions (e.g., “[hesitates]”) absent from natural patient speech. Each failure mode was addressed through targeted behavioural constraints appended to the prompt: an explicit safety block forbidding medical advice, a “one hidden item per turn” disclosure rule, conditional emotional response rules linking patient cooperation to interviewer empathy, and response length limits enforcing 1–3 sentence turns in colloquial register (representative prompt in Supplementary Table 2). The final prompts averaged 78 lines (∼1.8 KB) per scenario.

### 4.3 Platform and model selection

The AI-SP system was deployed as a web-based platform (Fig. 1) comprising a real-time chat interface, a scenario management module, an LLM integration layer routing messages to the assigned model, and an automated data collection backend recording all dialogue turns, timestamps, and session metadata. For Phase 2, a server-side whitelist access control mechanism enforced group allocation: upon login, each participant’s student ID was verified against the randomization list stored as a server environment variable, automatically assigning the correct study arm without self-selection. Only Arm A participants were granted access to the AI-SP chat interface; Arm B and Arm C participants received group-appropriate instructions but were technically unable to access the AI practice module, preventing cross-arm contamination at the platform level.

Five frontier LLMs were selected to span three accessibility tiers relevant to global deployment: **GPT-4o** (OpenAI; commercial API)[23], **Claude 4.5 Sonnet** (Anthropic; commercial API), **Gemini 2.5 Flash** (Google; free-tier API), **Qwen-2.5 Plus** (Alibaba; free-tier API), and **DeepSeek-R1** (DeepSeek; open-source, locally deployed). This selection enables comparisons across performance tiers (premium, cost-efficient, open-source), geographic provenance (US, China), and deployment models (cloud API, self-hosted), representing the full spectrum of options available to medical education programmes worldwide.

### 4.4 Phase 1: Dialogue generation and expert evaluation

For each scenario, two scripted student interaction patterns were developed representing distinct skill levels: competent (empathetic, open-ended, systematic) and novice (checklist-style, minimal empathy, jargon-heavy), based on documented communication patterns from institutional OSCE assessments[39]. Each scenario × student level × model combination was generated once using standardised parameters, yielding 50 dialogues (5 scenarios × 2 levels × 5 models).

Seven anaesthesiology faculty served as blinded evaluators (mean clinical experience: 12.4 years, range: 8–18; mean simulation education experience: 7.2 years). Evaluators were blinded to model identity and presentation order was randomised. Each evaluator assessed all 50 dialogues, yielding 350 total assessments. Six dimensions were scored on 1–5 Likert scales with anchored rubrics: Clinical Realism, Emotional Authenticity, Information Control, Communication Naturalness, Behavioural Consistency, and Pedagogical Value. Prior to scoring, evaluators completed a 3-hour calibration workshop. The educational viability threshold (≥20/30, equivalent to ≥3.33 mean per dimension) was established through a two-round modified Delphi process among the seven evaluators: the threshold corresponds to the “adequate for unsupervised student use” anchor point on the rubric, and maps to the institutional OSCE borderline pass standard (60% on a converted percentage scale) that the department applies to human SP encounters.

### 4.5 Phase 1b: Live student validation

Thirty-one medical students (fourth-and fifth-year) each completed all five scenarios using Qwen-2.5 Plus (selected as the deployment model because it achieved the highest Phase 1 fidelity score, offers a free-tier API with low latency in China, and natively handles Chinese-language interaction without translation artefacts), generating 155 live sessions. The platform recorded all dialogue turns, timestamps, and session duration in real time. Hidden-information discovery was assessed automatically: upon session completion, the dialogue transcript was submitted to a second Qwen-2.5 Plus instance with an analysis prompt listing all Layer 3 items for that scenario, returning a binary classification (discovered/not discovered) for each item. Each hidden item has explicit trigger conditions hard-coded in the system prompt (Supplementary Table 2), and the detection task reduces to verifying whether the patient character’s response contained the pre-specified disclosure content, a constrained binary classification with minimal ambiguity compared with open-ended assessment tasks. Following all scenarios, students completed an anonymous 11-item Likert-scale questionnaire (1–5) spanning AI-SP realism, learning value, and adoption willingness, plus three open-ended questions. Internal consistency was assessed using Cronbach’s *α*.

### 4.6 Phase 2: Randomized controlled trial

This phase was designed as a pilot RCT; the target sample size of approximately 60 was determined by the single-cohort rotation size available during the study period rather than a formal a priori power calculation, consistent with recommendations for pilot trials aimed at estimating effect sizes and assessing feasibility[26, 27]. Sixty-one medical students on anaesthesiology rotation were recruited. Participants were randomized using computer-generated sex-stratified block randomization (seed = 20260308; sex distribution: *χ*^2^ = 0.07, *p* = 0.956) into three arms: AI-SP (*n* = 22), human SP (*n* = 20), and control (*n* = 19). Three students withdrew before training commenced (one AI-SP, two human SP), yielding 58 analysed participants (AI-SP: 21, human SP: 18, control: 19; attrition rate: 4.9%). Students who had participated in Phase 1b were excluded to prevent contamination.

The entire protocol was designed as a fully remote intervention. Arm A (AI-SP) completed 10 practice consultations over one week (five scenarios, each attempted twice) using the text-based AI-SP platform, with automated post-session feedback. Arm B (Human SP) completed an identical schedule of 10 practice consultations via voice calls over videoconferencing software, with trained graduate student actors following identical scenario scripts and three-layer disclosure rules. The two arms intentionally used different communication modalities: text-based chat for AI-SP reflects the current deployment reality of LLM-driven systems, while voice interaction for human SP preserves the ecological validity of actor-based training[34]. This design prioritises external validity, comparing AI-SP and human SP as they would actually be deployed, over strict internal control of modality. The implications of this modality difference are addressed in the Limitations. Arm C (Control) followed the standard curriculum with no additional practice. To prevent cross-arm contamination, the AI-SP platform enforced server-side whitelist access control: only Arm A student identifiers were authorised for the practice module.

The OSCE comprised two scoring instruments: a 14-item binary checklist assessing observable consultation behaviours (e.g., “verified patient identity”, “explained anaesthetic options”) and a 5-item global rating scale assessing holistic communication quality on 1–5 Likert scales with descriptive anchors (e.g., 1 = “no attempt”, 3 = “adequate but formulaic”, 5 = “exemplary, patient-centred”). The primary outcome was pre–post change in the OSCE checklist score (/14), selected a priori based on its superior inter-rater reliability (ICC(2,1) = 0.72) compared with the global rating scale (ICC = 0.15) and total score (ICC = 0.40). The poor global rating ICC likely reflects the inherently subjective nature of holistic quality judgements in asynchronous video scoring without real-time calibration; leave-one-out and sensitivity analyses (Supplementary Note 1, Supplementary Figs. 6) confirmed that this was not driven by a single outlier examiner but reflected systematic inter-examiner variability across all five dimensions. OSCEs were conducted at baseline (Week 0) and immediately post-training (Week 1) across five stations (10 minutes each) via remote videoconference. All sessions were video-recorded and scored asynchronously by five blinded examiners, each assigned primary responsibility for one station with cross-scoring of 15 randomly selected recordings per station for ICC estimation (26% dual-rated). This video-recorded OSCE approach is consistent with validated telehealth assessment protocols[34, 35]. Secondary outcomes included total OSCE score change (/39), self-efficacy change (10-item questionnaire, /50; assessed at Week 0 and Week 3), and training satisfaction (11-item Likert scale, /55; collected for Arms A and B only, as Arm C received no intervention to evaluate).

### 4.7 Statistical analysis

Phase 1 analyses employed one-way ANOVA (five models) and two-way ANOVA (model × student level; model × complexity) with Bonferroni-corrected post-hoc comparisons. Inter-rater reliability was assessed using ICC (two-way random, absolute agreement). Effect sizes are reported as *η*^2^ (ANOVA) and Cohen’s *d* (pairwise).

Phase 1b used descriptive statistics for per-scenario and per-item discovery rates. Sequential learning effects were assessed using paired *t*-tests (S1 vs S5). Pearson correlation assessed the engagement–performance relationship.

Phase 2 employed non-parametric tests given the small sample and non-normal distributions. Between-group comparisons used Kruskal-Wallis H tests with pairwise Mann-Whitney U tests. Within-group pre–post comparisons used Wilcoxon signed-rank tests. ANCOVA (post-training score ∼ group + baseline score) was used to adjust for baseline differences. Effect sizes are reported as Cohen’s *d* (between-group) and paired *d* (within-group). Inter-rater reliability was assessed using ICC(2,1) (two-way random, single measures, absolute agreement). All analyses were intention-to-treat. Statistical significance: *α* = 0.05. Analyses used Python 3.9 (SciPy 1.11, statsmodels 0.14).

## Data Availability

Anonymised data and analysis code generated during this study will be made available from the corresponding author on reasonable request. The AI-SP platform code and three-layer information architecture specifications can be shared subject to a standard collaboration agreement.

## Acknowledgements

The authors thank the expert evaluators for their contributions to the fidelity assessment, the graduate student standardised patient actors in Phase 2, and all 92 participating medical students from Dalian Medical University.

## Declarations

### Author contributions

P.W. conceptualised the study, designed the methodology and AI-SP prompts, curated the data, performed the formal analysis and visualisation, and wrote the original draft. Y.H. contributed to formal analysis, visualisation, and software development, and reviewed and edited the manuscript. J.Z., Y.L., and M.J. contributed to investigation and validation. X.L., H.Z., D.X., H.M., and L.W. contributed to investigation through OSCE examination and scoring. Q.W. conceptualised the study, designed the methodology, supervised the project, administered the project, acquired funding, and reviewed and edited the manuscript. All authors reviewed and approved the final manuscript.

### Data availability

Anonymised evaluation scores, AI-SP prompt templates, scenario specifications, scoring rubrics, representative dialogue transcripts, and the DECIDE-AI reporting checklist are available at: https://github.com/RegAItool/ai-standardized-patient. Source data for all figures and tables are provided with the paper.

### Code availability

Analysis scripts (Python 3.9) for all statistical analyses and figure generation are available at: https://github.com/RegAItool/ai-standardized-patient.

### Ethics approval

The study was determined to be exempt from formal ethics review by the Ethics Committee of The First Affiliated Hospital of Dalian Medical University (exemption reference: YJ-MS-2025-02; document ID: AF/SW-31/2023-01.0), as it involved comparison of standard educational methods among medical students with no patient contact, no clinical intervention, and no collection of biological specimens. All student participants provided written informed consent. Participation was voluntary; non-participation did not affect academic standing.

### Reporting summary

Further information on research design is available in the Nature Portfolio Reporting Summary linked to this article.

## Supplementary Information

Supplementary Information accompanies this paper and includes: Supplementary Table 1 (DECIDE-AI reporting checklist), Supplementary Table 2 (complete AI-SP prompt templates for all five scenarios), Supplementary Table 3 (per-scenario, per-evaluator fidelity scores), Supplementary Table 4 (per-scenario OSCE checklist analysis), Supplementary Table 5 (self-efficacy per-item analysis), Supplementary Table 6 (CONSORT checklist), Supplementary Tables 7–8 (ICC by scoring instrument and examiner patterns), Supplementary Tables 9–10 (global rating ICC robustness: leave-one-out and sensitivity analyses), Supplementary Note 1 (global rating ICC robustness analysis), Supplementary Figs. 1–4 (per-dimension fidelity, OSCE trajectories, self-efficacy radar, satisfaction comparison), Supplementary Fig. 5 (ICC robustness: checklist–global correlation, sensitivity analysis, examiner variability), Supplementary Fig. 6 (iceberg model of three-layer information architecture), and Supplementary Data (complete anonymised dialogue transcripts and evaluation scores).

## References

[1] Makoul, G. Essential elements of communication in medical encounters: The Kalamazoo consensus statement. Academic Medicine 76, 390–393 (2001).

[2] Ericsson, K. A. Deliberate practice and the acquisition and maintenance of expert performance in medicine and related domains. Academic Medicine 79, S70–S81 (2004).

[3] Barrows, H. S. An overview of the uses of standardized patients for teaching and evaluating clinical skills. Academic Medicine 68, 443–451 (1993).

[4] Issenberg, S. B., McGaghie, W. C., Petrusa, E. R., Gordon, D. L. & Scalese, R. J. Features and uses of high-fidelity medical simulations that lead to effective learning: A BEME systematic review. Medical Teacher 27, 10–28 (2005).

[5] Blitz, J. D., Kendale, S. M., Jain, S. K. et al. Preoperative evaluation clinic visit is associated with decreased risk of in-hospital postoperative mortality. Anesthesiology 125, 280–294 (2016).

[6] Bosse, H. M., Nickel, M., Huwendiek, S., Schultz, J. H. & Nikendei, C. Cost-effectiveness of peer role play and standardized patients in undergraduate communication training. BMC Medical Education 15, 183 (2015).

[7] Singhal, K., Azizi, S., Tu, T. et al. Large language models encode clinical knowledge. Nature 620, 172–180 (2023).

[8] Savage, T., Nayak, A., Gallo, R., Rangan, E. & Chen, J. H. Diagnostic reasoning prompts reveal the potential for large language model interpretability in medicine. npj Digital Medicine 7, 20 (2024).

[9] Lee, P., Bubeck, S. & Petro, J. Benefits, limits, and risks of GPT-4 as an AI chatbot for medicine. New England Journal of Medicine 388, 1233–1239 (2023).

[10] Shanahan, M., McDonell, K. & Reynolds, L. Role play with large language models. Nature 623, 493–498 (2023).

[11] Kron, F. W., Fetters, M. D., Scerbo, M. W. et al. Using a computer simulation for teaching communication skills: A blinded multisite mixed methods randomized controlled trial. Patient Education and Counseling 100, 748–759 (2017).

[12] Lane, H. C., Hays, M. J., Core, M. G. & Auerbach, D. Learning intercultural communication skills with virtual humans: Feedback and fidelity. Journal of Educational Psychology 107, 254–270 (2015).

[13] Thesen, T. & Park, S. H. A generative AI teaching assistant for personalized learning in medical education. npj Digital Medicine 8, 627 (2025).

[14] Mandal, S., Wiesenfeld, B. M., Szerencsy, A. C. et al. Utilization of generative AI-drafted responses for managing patient-provider communication. npj Digital Medicine 8, 591 (2025).

[15] Stanceski, K., Zhong, S., Zhang, X. et al. The quality and safety of using generative AI to produce patient-centred discharge instructions. npj Digital Medicine 7, 329 (2024).

[16] Luo, M.-J. et al. A large language model digital patient system enhances ophthalmology history taking skills. npj Digital Medicine 8, 502 (2025).

[17] Schwill, S., et al. AI-standardized clinical examination training on OSCE performance. NEJM AI 2 (2025).

[18] Yu, H., Zhou, J., Li, L. et al. Simulated patient systems powered by large language model-based AI agents offer potential for transforming medical education. Communications Medicine 6, 27 (2025).

[19] Haltaufderheide, J. & Ranisch, R. The ethics of ChatGPT in medicine and health-care: a systematic review on large language models. npj Digital Medicine 7, 183 (2024).

[20] Nestel, D. & Tierney, T. Role-play for medical students learning about communication: Guidelines for maximising benefits. BMC Medical Education 7, 3 (2007).

[21] Collins, A., Brown, J. S. & Holum, A. Cognitive apprenticeship: Making thinking visible. American Educator 15, 6–11 (1991).

[22] Zhang, G. et al. Closing the gap between open source and commercial large language models for medical evidence summarization. npj Digital Medicine 7, 239 (2024).

[23] Wu, C., Qiu, P., Liu, J. et al. Towards evaluating and building versatile large language models for medicine. npj Digital Medicine 8, 58 (2025).

[24] Jin, Q. et al. Hidden flaws behind expert-level accuracy of multimodal GPT-4 vision in medicine. npj Digital Medicine 7, 190 (2024).

[25] McGaghie, W. C., Issenberg, S. B., Cohen, E. R., Barsuk, J. H. & Wayne, D. B. Does simulation-based medical education with deliberate practice yield better results than traditional clinical education? a meta-analytic comparative review of the evidence. Academic Medicine 86, 706–711 (2011).

[26] Lancaster, G. A., Dodd, S. & Williamson, P. R. Design and analysis of pilot studies: Recommendations for good practice. Journal of Evaluation in Clinical Practice 10, 307–312 (2004).

[27] Thabane, L. et al. A tutorial on pilot studies: The what, why, and how. BMC Medical Research Methodology 10, 1 (2010).

[28] Faul, F., Erdfelder, E., Lang, A.-G. & Buchner, A. G*Power 3: A flexible statistical power analysis program for the social, behavioral, and biomedical sciences. Behavior Research Methods 39, 175–191 (2007).

[29] Bandura, A. Self-efficacy: Toward a unifying theory of behavioral change. Psychological Review 84, 191–215 (1977).

[30] Mehrabian, A. Silent Messages: Implicit Communication of Emotions and Attitudes (Wadsworth, 1971).

[31] Walther, J. B. Computer-mediated communication: Impersonal, interpersonal, and hyperpersonal interaction. Communication Research 23, 3–43 (1996).

[32] Regehr, G., MacRae, H., Reznick, R. K. & Szalay, D. Comparing the psychometric properties of checklists and global rating scales for assessing performance on an OSCE-format examination. Academic Medicine 73, 993–997 (1998).

[33] Gershov, S., Mahameed, F., Raz, A. & Laufer, S. Towards accurate and interpretable competency-based assessment: Enhancing clinical competency assessment through multimodal AI and anomaly detection. npj Digital Medicine 9, 219 (2026).

[34] Boursicot, K. et al. Conducting a high-stakes OSCE in a COVID-19 environment. MedEdPublish 9, 54 (2020).

[35] Hopwood, J., Myers, G. & Sturrock, A. Remote OSCE experience: What first year medical students thought. BMC Medical Education 22, 363 (2022).

[36] Holderried, F. et al. Large language models improve clinical decision making of medical students through patient simulation and structured feedback. BMC Medical Education 24, 1391 (2024).

[37] Vasey, B. et al. Reporting guideline for the early-stage clinical evaluation of decision support systems driven by artificial intelligence: DECIDE-AI. Nature Medicine 28, 924–933 (2022).

[38] Kurtz, S., Silverman, J. & Draper, J. Teaching and learning communication skills in medicine (2003).

[39] Frank, J. R., Snell, L. S., ten Cate, O. et al. Competency-based medical education: Theory to practice. Medical Teacher 32, 638–645 (2010).

