## Supplementary Information for "From simulation to pedagogy: structured AI standardized patients for clinical communication training validated through multi-model and randomized evaluation"

#### Supplementary Table 1: DECIDE-AI reporting checklist

| # | Item | Reported as | Location |
|---|---|---|---|
| **Title and abstract** | | | |
| 1 | Title clearly identifies the study as an evaluation of an AI-based decision support system | Title specifies "AI standardized patients" and evaluation design | Title |
| 2 | Abstract describes the AI system, evaluation design, main results and conclusions | Structured abstract covers AI-SP system, three-phase design, quantitative results and implications | Abstract |
| **Background and objectives** | | | |
| 3 | Scientific and clinical background | Limitations of human SP programmes, role of LLMs in simulation training | Introduction |
| 4 | Primary objective of evaluation | Three objectives stated: fidelity validation, ecological validation, RCT efficacy | Introduction |
| **Study design** | | | |
| 5 | Type of evaluation study | Three-phase design: blinded expert evaluation (Phase 1), live validation (Phase 1b), three-arm RCT (Phase 2) | Methods 4.1 |
| 6 | Ethical approval and participant consent | Ethics approval DLMU-2024-AE-012 (Phases 1/1b); Phase 2 exempt; written informed consent obtained | Methods 4.1 |
| **Participants** | | | |
| 7 | Eligibility criteria for participants | Phase 1: 7 anaesthesiology faculty (≥8 yrs experience); Phase 1b: 4th/5th-year medical students; Phase 2: students on anaesthesiology rotation | Methods 4.4–4.6 |
| 8 | Setting and location | Single-institution; Department of Anaesthesiology, The First Affiliated Hospital of Dalian Medical University | Methods 4.1 |

*(continued)*

| # | Item | Reported as | Location |
|---|---|---|---|
| **AI system description** | | | |
| 9 | Description of AI system, including intended use and target population | Three-layer information architecture; five frontier LLMs evaluated; intended for preoperative communication training | Introduction, Methods 4.2–4.3 |
| 10 | AI system development and training | Prompt engineering with iterative expert consensus; standardised parameters across models | Methods 4.2 |
| 11 | Version and model identifiers | GPT-4o, Claude 4.5 Sonnet (claude-sonnet-4-5-20250929), Gemini 2.5 Flash, Qwen-2.5 Plus, DeepSeek-R1 | Methods 4.3 |
| **Outcomes** | | | |
| 12 | Primary and secondary outcome measures | Primary: OSCE checklist score change; secondary: total OSCE score, self-efficacy, satisfaction | Methods 4.6 |
| 13 | Outcome measurement methods and timing | Blinded asynchronous video scoring; dual-rating of 26% for ICC; self-efficacy at Weeks 0 and 3 | Methods 4.6 |
| **Statistical analysis** | | | |
| 14 | Statistical methods used | ANOVA, ICC, Kruskal-Wallis, Wilcoxon, ANCOVA; effect sizes ($\eta^2$, Cohen's $d$) | Methods 4.7 |
| 15 | Sample size justification | Pilot sizing; post-hoc power reported ($N \geq 216$ for 80% power) | Methods 4.6, Discussion |
| **Results** | | | |
| 16 | Participant flow and numbers analysed | 61 randomised, 3 withdrew, 58 analysed (ITT); CONSORT flow diagram provided | Results 2.3, Fig. 4 |
| 17 | Baseline characteristics | Group comparability at baseline confirmed (all $p > 0.19$) | Results 2.3, Table 3 |
| 18 | Main results with effect sizes and CIs | OSCE: $p = 0.483$, $d_{AB} = 0.38$; self-efficacy: $p = 0.034$, $d = 0.62$; satisfaction: $p = 0.876$ | Results 2.3, Table 3 |
| 19 | Harms and unintended consequences | No adverse events; cross-arm contamination prevented by server-side whitelist | Methods 4.3, Results 2.3 |
| **Discussion** | | | |
| 20 | Limitations | Sample size, testing effect, single institution, modality difference, immediate outcomes only | Discussion 3.4 |

*(continued)*

| # | Item | Reported as | Location |
|---|---|---|---|
| 21 | Generalisability | Three-layer architecture is model-agnostic and domain-agnostic; single-centre Chinese-language context limits external validity | Discussion 3.4 |
| 22 | Overall conclusions consistent with evidence | Conclusions supported by multi-phase validation; limitations explicitly acknowledged | Discussion |
| **Other information** | | | |
| 23 | Data and code availability | Anonymised data and analysis scripts available at GitHub | Data availability |
| 24 | Funding and competing interests | Declared; no competing interests | Declarations |

**Supplementary Table 1.** DECIDE-AI reporting checklist (Vasey et al., 2022, *BMJ*). Items are mapped to the corresponding section of the manuscript.

#### Supplementary Table 3: Per-scenario, per-evaluator fidelity scores

**Supplementary Table 3a.** Mean total fidelity score (out of 30) by evaluator and scenario. Each cell represents mean across 10 dialogues (5 models × 2 student skill levels).

| Evaluator | S1 | S2 | S3 | S4 | S5 | Overall |
|---|---|---|---|---|---|---|
| Evaluator 1 | 20.4 | 19.9 | 20.7 | 18.7 | 19.0 | 19.7 |
| Evaluator 2 | 18.8 | 18.2 | 18.2 | 19.0 | 20.2 | 18.9 |
| Evaluator 3 | 20.6 | 19.0 | 19.2 | 19.4 | 18.8 | 19.4 |
| Evaluator 4 | 23.0 | 21.1 | 21.0 | 21.6 | 21.5 | 21.6 |
| Evaluator 5 | 19.8 | 19.8 | 18.8 | 19.4 | 19.5 | 19.5 |
| Evaluator 6 | 21.6 | 19.1 | 20.0 | 18.6 | 18.8 | 19.6 |
| Evaluator 7 | 21.5 | 21.3 | 22.9 | 21.8 | 20.8 | 21.7 |
| **Grand mean** | **20.8** | **19.9** | **20.1** | **19.8** | **19.8** | **20.1** |

**Supplementary Table 3b.** Mean score per evaluation dimension by evaluator (scale 1–5; averaged across all 50 dialogues). D1 = Clinical Realism; D2 = Emotional Authenticity; D3 = Information Control; D4 = Communication Naturalness; D5 = Behavioural Consistency; D6 = Pedagogical Value.

| Evaluator | D1 | D2 | D3 | D4 | D5 | D6 |
|---|---|---|---|---|---|---|
| Evaluator 1 | 3.18 | 2.96 | 3.32 | 3.24 | 3.42 | 3.62 |
| Evaluator 2 | 3.10 | 3.16 | 2.90 | 3.26 | 3.36 | 3.10 |
| Evaluator 3 | 3.00 | 3.12 | 3.28 | 3.32 | 3.32 | 3.36 |
| Evaluator 4 | 3.68 | 3.54 | 3.32 | 3.54 | 4.00 | 3.56 |
| Evaluator 5 | 3.18 | 3.34 | 3.28 | 3.48 | 3.34 | 2.84 |
| Evaluator 6 | 3.52 | 3.50 | 3.08 | 3.22 | 3.32 | 2.98 |
| Evaluator 7 | 4.00 | 3.82 | 3.56 | 3.44 | 3.48 | 3.36 |
| **Grand mean** | **3.38** | **3.35** | **3.25** | **3.36** | **3.46** | **3.26** |

**Inter-evaluator consistency.** The educational viability threshold (≥20/30) was exceeded by 4 of 7 evaluators across all five scenarios. Evaluator 2 rated below threshold in three scenarios (S1–S3), reflecting a consistently more stringent standard rather than a scenario-specific artefact. The ICC(2,k) = 0.794 for the seven-evaluator panel indicates good inter-rater reliability at the panel level.

#### Supplementary Figure 1: Per-dimension fidelity scores by model and student skill level

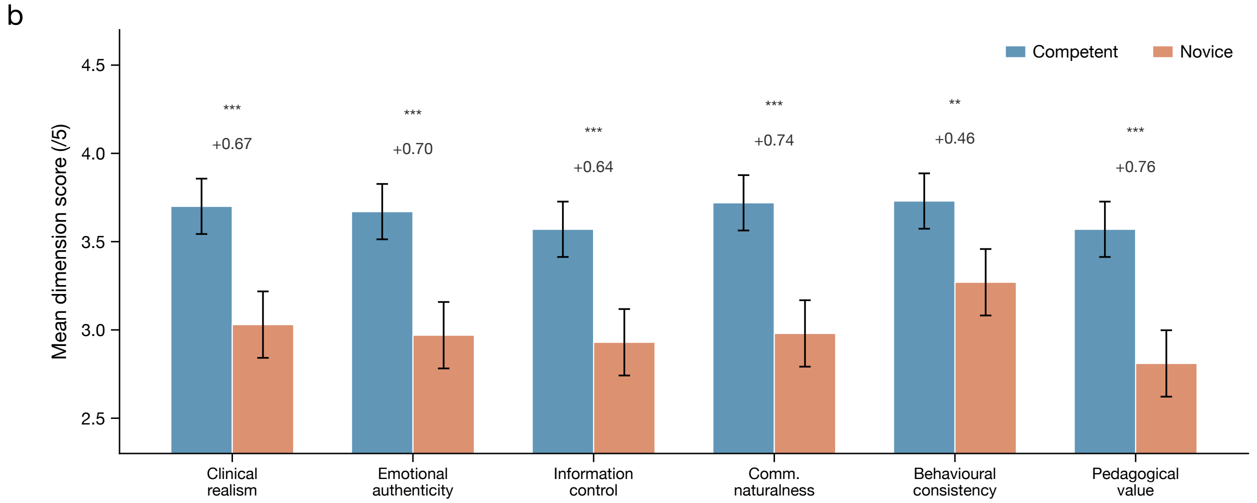


**Supplementary Figure 1.** Fidelity scores across six evaluation dimensions by model, separated by student skill level (competent vs novice). Each panel shows mean ± SD across seven evaluators and five scenarios. The competent–novice gap is consistent across all six dimensions and all models, confirming that the three-layer architecture reliably operationalises student skill differentiation regardless of the underlying LLM. Models are colour-coded by accessibility tier: commercial premium (GPT-4o, Claude 4.5 Sonnet), cost-efficient (Gemini 2.5 Flash, Qwen-2.5 Plus), and open-source (DeepSeek-R1).

### Supplementary Figure 2: OSCE checklist score trajectories by scenario and study arm

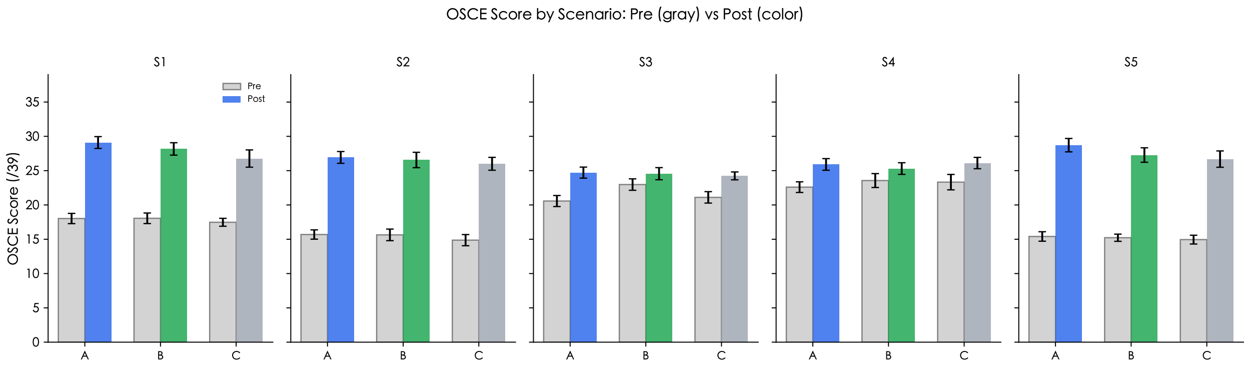


**Supplementary Figure 2.** Pre- to post-training OSCE checklist score trajectories by scenario (S1–S5) and study arm. Each point shows group mean; error bars are ± SEM. The largest gains occurred in S5 (mastectomy) and S2 (thyroidectomy) across all arms. S4 (lumbar disc) showed minimal change, consistent with ceiling effects at baseline (mean 8.2–8.5/14). No scenario showed a statistically significant between-arm difference. Data based on complete scored records ($n$ = 21/18/19 per arm for all scenarios; full dataset from primary scoring plus cross-rating.

#### Supplementary Table 6: CONSORT 2010 reporting checklist (Phase 2 RCT)

| # | Item | Reported as | Location |
|---|---|---|---|
| **Title and abstract** | | | |
| 1a | Identification as a randomized trial in title | "randomized evaluation" in title | Title |
| 1b | Structured summary of design, methods, results, conclusions | Included; RCT described alongside Phase 1/1b | Abstract |
| **Introduction** | | | |
| 2a | Scientific background and explanation of rationale | Gap in AI-SP evidence and need for human SP comparator established | Introduction |
| 2b | Specific objectives or hypotheses | Three research questions stated for Phase 2 | Results 2.3 |
| **Methods — Participants** | | | |
| 3a | Eligibility criteria for participants | Medical students on anaesthesiology rotation; Phase 1b participants excluded | Methods 4.6 |
| 3b | Settings and locations where data were collected | Remote protocol; single institution; Department of Anaesthesiology, DLMU | Methods 4.1 |
| **Methods — Interventions** | | | |
| 4 | Interventions for each group; sufficient detail for replication | Arm A: 10 AI-SP sessions (text, 1 week); Arm B: 10 voice SP sessions (identical scripts); Arm C: standard curriculum | Methods 4.6 |
| **Methods — Outcomes** | | | |
| 5 | Pre-specified primary and secondary outcomes, and when assessed | Primary: OSCE checklist score change (Week 0 to Week 1); secondary: total score, self-efficacy (Week 3), satisfaction | Methods 4.6 |
| **Methods — Sample size** | | | |
| 6 | Sample size determination | Pilot study; post-hoc power reported ($N \geq 216$ for 80% power, $d = 0.38$) | Discussion 3.3 |
| **Methods — Randomization** | | | |
| 7a | Sequence generation method | Computer-generated sex-stratified block randomization (seed = 20260308) | Methods 4.6 |
| 7b | Type of randomization; details of any restriction | Stratified block; sex as stratification variable ($\chi^2 = 0.07$, $p = 0.956$) | Methods 4.6 |
| 8 | Allocation concealment mechanism | Server-side whitelist enforced arm assignment; students could not self-select | Methods 4.3 |

*(continued)*

| # | Item | Reported as | Location |
|---|---|---|---|
| 9 | Implementation (who generated sequence, enrolled, assigned) | Allocation list generated by P.W.; assignments enforced automatically by platform | Methods 4.3 |
| **Methods — Blinding** | | | |
| 10 | Blinding details | OSCE examiners blinded to arm allocation; students not blinded (inherent to intervention design) | Methods 4.6 |
| **Methods — Statistical methods** | | | |
| 11 | Statistical methods for primary outcome and additional analyses | Kruskal-Wallis, Mann-Whitney, Wilcoxon, ANCOVA; effect sizes; ICC | Methods 4.7 |
| **Results — Participant flow** | | | |
| 12a | Numbers randomized, receiving intended treatment, analysed | 61 randomised; 58 analysed (3 withdrew before training) | Results 2.3, Fig. 4 |
| 12b | Losses and exclusions after randomization | 1 (Arm A), 2 (Arm B) withdrew; 0 excluded post-allocation | Results 2.3, Fig. 4 |
| **Results — Recruitment** | | | |
| 13 | Dates defining recruitment periods | March 2026 (anaesthesiology rotation cohort) | Methods 4.6 |
| **Results — Baseline data** | | | |
| 14 | Baseline demographic and clinical characteristics | OSCE checklist, total score, self-efficacy comparable across arms | Results 2.3, Table 3 |
| **Results — Numbers analysed** | | | |
| 15 | Number in each group included in each analysis | All 58 analysed (intention-to-treat); no imputation required | Results 2.3 |
| **Results — Outcomes and estimation** | | | |
| 16 | Results for each outcome: effect size and precision | Checklist: KW $p = 0.483$, $d_{AB} = 0.38$; self-efficacy: $p = 0.034$, $d = 0.62$; satisfaction: $p = 0.876$ | Results 2.3, Table 3 |
| **Results — Ancillary analyses** | | | |
| 17 | Results of any other analyses (subgroup, sensitivity) | ANCOVA adjusting for baseline; three scoring instruments (sensitivity); per-scenario analysis | S-Table 4, S-Table 10 |
| **Results — Harms** | | | |
| 18 | All important harms or unintended effects | No adverse events; cross-arm contamination prevented | Methods 4.3 |
| **Discussion** | | | |

*(continued)*

| # | Item | Reported as | Location |
|---|---|---|---|
| 19 | Trial limitations | Power, testing effect, modality difference, single institution, immediate outcomes | Discussion 3.4 |
| 20 | Generalisability | Architecture is model-agnostic; Chinese-language, single centre limits external validity | Discussion 3.4 |
| 21 | Interpretation consistent with results | Equivalence inference contextualised within pilot limitations | Discussion 3.3 |
| **Other information** | | | |
| 22 | Registration | Educational pilot; formal registration not required by institutional ethics committee | Methods 4.1 |
| 23 | Protocol | Study protocol available from corresponding author on reasonable request | — |
| 24 | Funding | Declared in Acknowledgements | Declarations |

**Supplementary Table 6.** CONSORT 2010 checklist for reporting parallel-group randomized trials. Items correspond to the Phase 2 three-arm RCT component of this study.

#### Supplementary Table 7: Inter-rater reliability by scoring instrument and scenario

**Supplementary Table 7a.** ICC(2,1) by scoring instrument (OSCE-1, $n = 146$ dual-rated pairs).

| Scoring instrument | ICC(2,1) | 95% CI | Interpretation |
|---|---|---|---|
| Checklist (14 binary items, /14) | 0.717 | [0.628, 0.788] | Moderate–Good |
| Global rating (5 Likert items, /25) | 0.146 | [0.040, 0.351] | Poor |
| Total score (/39) | 0.400 | [0.335, 0.588] | Poor |

**Supplementary Table 7b.** Checklist ICC(2,1) by scenario (OSCE-1).

| Scenario | ICC(2,1) | 95% CI | $n$ pairs |
|---|---|---|---|
| S1 Cholecystectomy | 0.580 | [0.386, 0.731] | 58 |
| S2 Thyroidectomy | 0.843 | [0.690, 0.974] | 11 |
| S3 Knee replacement | 0.644 | [0.149, 0.871] | 13 |
| S4 Lumbar disc | 0.778 | [0.659, 0.871] | 54 |
| S5 Mastectomy | 0.720 | [0.246, 0.928] | 10 |

**Supplementary Table 7c.** Score disagreement between dual-rated examiners (OSCE-1, $n = 146$ pairs).

| Metric | Checklist (/14) | Global (/25) | Total (/39) |
|---|---|---|---|
| Mean absolute difference | 1.2 | 4.8 | 5.1 |
| Exact agreement (%) | 27% | 6% | 5% |
| Difference $\leq 2$ (%) | 91% | 29% | 26% |
| Difference $> 5$ (%) | 0% | 42% | 46% |
| Maximum difference | 4 | 13 | 15 |

**Rationale for primary outcome selection.** The 14-item binary checklist achieved ICC(2,1) = 0.717, classified as moderate-to-good reliability. The 5-item global rating scale showed poor reliability (ICC = 0.146), with 42% of dual-rated pairs differing by more than 5 points on a 25-point scale. This disparity reflects the inherent subjectivity of holistic performance judgments versus binary behavioural checklist items, consistent with prior OSCE reliability literature. The checklist score was therefore designated as the primary outcome measure.

**Examiner scoring patterns.** Analysis of global rating means by examiner revealed systematic calibration differences: two examiners scored substantially lower (mean 6.7 and 7.6/25) than the remaining three (mean 13.4–15.9/25), while checklist scores were comparable across all five examiners (7.3–7.9/14). This pattern indicates consistent application of objective checklist criteria alongside divergent subjective standards, further supporting the checklist as the more reliable measure.

#### Supplementary Table 5: Self-efficacy per-item change

**Supplementary Table 5.** Self-efficacy change (Δ) by item and group (Week 0 → Week 3).

| Item | AI-SP Δ | Human SP Δ | Control Δ | $p$ (KW) |
|---|---|---|---|---|
| 1. History taking | +1.10 | +0.83 | +0.47 | 0.094 |
| 2. Eliciting hidden information | +1.10 | +0.67 | +0.74 | 0.183 |
| 3. Open-ended questioning | +1.10 | +1.00 | +0.63 | 0.208 |
| 4. Emotional response | +0.81 | +0.56 | +0.84 | 0.380 |
| 5. Explaining anaesthesia | +0.86 | +0.56 | +0.37 | 0.165 |
| 6. Handling patient defenses | +1.00 | +0.50 | +0.89 | 0.247 |
| 7. Managing time pressure | +1.10 | +0.50 | +0.53 | 0.060 |
| 8. Handling special cases | +1.24 | +0.83 | +0.74 | 0.277 |
| 9. Building trust | +0.95 | +0.78 | +0.47 | 0.275 |
| 10. Overall confidence | +0.95 | +0.61 | +0.42 | 0.140 |
| **Total (/50)** | **+10.2** | **+6.8** | **+6.1** | **0.064** |

AI-SP training produced the largest gains across all 10 items. Item 7 (managing time pressure) approached significance ($p = 0.060$), with AI-SP students improving more than twice as much as human SP or control students. This may reflect the self-paced nature of AI-SP practice, which allows students to develop time management strategies without the social pressure of a real-time interlocutor.

#### Supplementary Table 4: Per-scenario OSCE checklist analysis

**Supplementary Table 4.** OSCE checklist score change by scenario and group.

| Scenario | Group | $n$ | Pre | Post | Δ | $p$ (Wilcoxon) |
|---|---|---|---|---|---|---|
| S1 Cholecystectomy | AI-SP | 21 | 7.6 | 8.8 | +1.2 | 0.008 |
| | Human SP | 18 | 7.3 | 7.9 | +0.6 | 0.080 |
| | Control | 19 | 7.1 | 7.6 | +0.5 | 0.350 |
| | Δ between-group: $p = 0.599$ | | | | | |
| S2 Thyroidectomy | AI-SP | 21 | 8.2 | 9.4 | +1.2 | 0.011 |
| | Human SP | 18 | 8.0 | 9.7 | +1.7 | 0.006 |
| | Control | 19 | 7.7 | 9.1 | +1.4 | 0.010 |
| | Δ between-group: $p = 0.898$ | | | | | |
| S3 Knee replacement | AI-SP | 21 | 6.1 | 7.2 | +1.1 | 0.093 |
| | Human SP | 18 | 7.4 | 7.3 | −0.1 | 0.756 |
| | Control | 19 | 6.0 | 6.7 | +0.7 | 0.091 |
| | Δ between-group: $p = 0.392$ | | | | | |
| S4 Lumbar disc | AI-SP | 21 | 8.2 | 8.6 | +0.4 | 0.412 |
| | Human SP | 18 | 8.5 | 8.3 | −0.2 | 0.635 |
| | Control | 19 | 8.4 | 8.4 | +0.0 | 0.965 |
| | Δ between-group: $p = 0.662$ | | | | | |
| S5 Mastectomy | AI-SP | 21 | 7.4 | 10.3 | +2.9 | <0.001 |
| | Human SP | 18 | 7.2 | 9.9 | +2.7 | 0.001 |
| | Control | 19 | 7.1 | 9.3 | +2.2 | 0.005 |
| | Δ between-group: $p = 0.497$ | | | | | |

Improvements were largest in S5 (mastectomy) across all groups; S2 (thyroidectomy) also showed consistent significant improvement across all arms ($p < 0.02$). S4 (lumbar disc) showed the smallest gains, reflecting a ceiling effect (baseline scores 8.2–8.5/14). S3 (knee replacement) showed no significant within-group improvement in any arm, suggesting its checklist items were not well-targeted by the one-week training protocol. No individual scenario showed a significant between-group difference in Δ scores.

#### Supplementary Figure 3: Self-efficacy radar plots

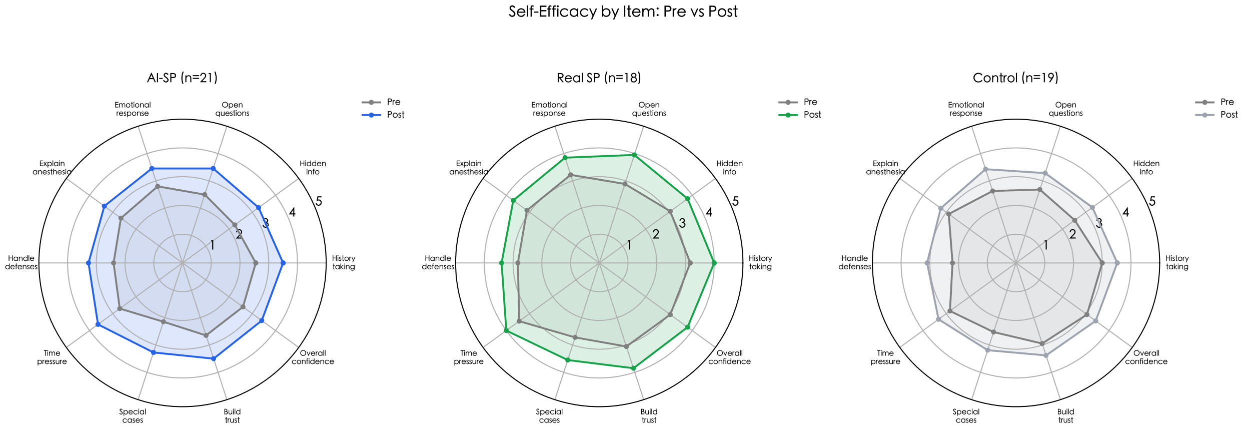


**Supplementary Figure 3.** Self-efficacy scores by item (1–5 Likert scale) at pre-training (gray) and post-training (colour) for each study arm. The AI-SP group (blue) shows the most uniform expansion across all ten dimensions, with the largest gains in managing time pressure (item 7) and handling special cases (item 8).

#### Supplementary Figure 4: Training satisfaction comparison

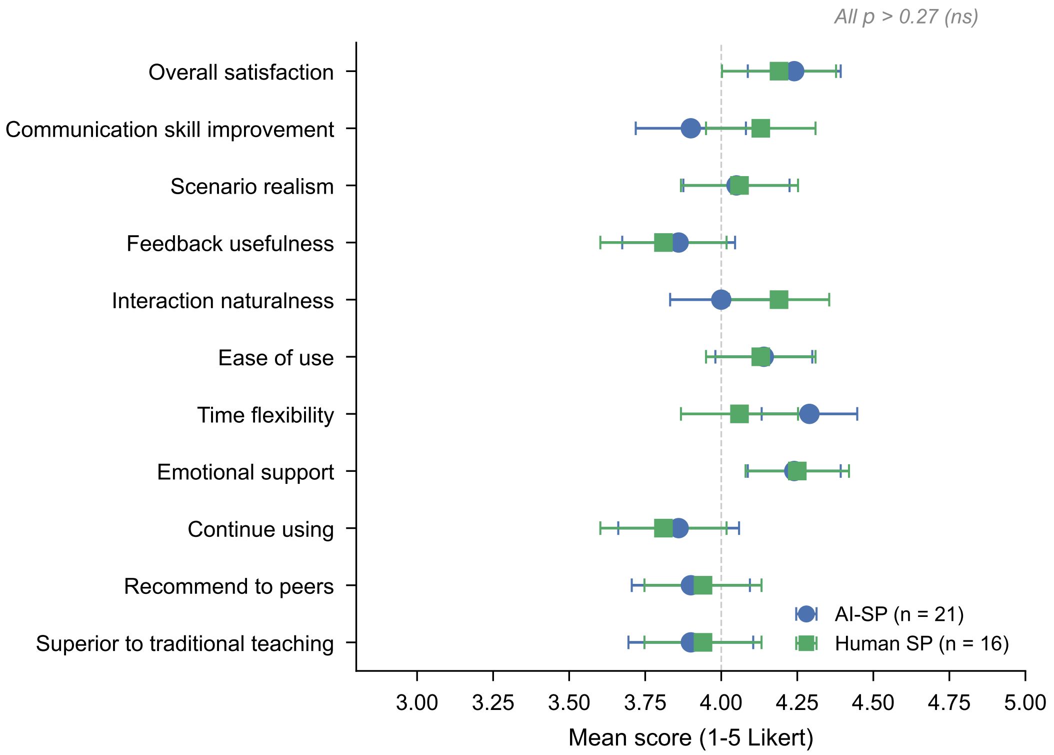


**Supplementary Figure 4.** Training satisfaction scores (1–5 Likert) across 11 dimensions for AI-SP ($n = 21$) and Human SP ($n = 16$) arms. No dimension showed a significant between-arm difference (all Mann-Whitney $p > 0.29$). Both arms scored above 3.8 on all items.

#### Supplementary Table 8: Examiner scoring patterns

**Supplementary Table 8.** Mean scores by examiner (OSCE-1), illustrating divergent global rating calibration.

| Examiner | $n$ records | Checklist mean (/14) | Global mean (/25) | Total mean (/39) |
|---|---|---|---|---|
| Zhang Haibin | 70 | 7.8 | 6.7 | 14.5 |
| Wang Lihong | 117 | 7.3 | 7.6 | 15.0 |
| Lu Xinyu | 69 | 7.3 | 13.4 | 20.7 |
| Xu Danyang | 69 | 7.9 | 13.6 | 21.5 |
| Ming Hao | 120 | 7.5 | 15.9 | 23.4 |

All five examiners scored checklist items within a narrow range (7.3–7.9/14, range = 0.6), confirming consistent application of binary behavioural criteria. In contrast, global ratings spanned a 2.4-fold range (6.7–15.9/25), with two examiners (Zhang Haibin, Wang Lihong) scoring at approximately 30% of maximum and three examiners scoring at 53–64%. This systematic calibration difference, rather than random disagreement, accounts for the low global rating ICC. The dual-rating design (26% of student-scenario pairs cross-rated) enables detection and transparent reporting of this examiner effect without compromising the validity of the primary (checklist-based) analysis.

#### Supplementary Note 1: Global rating ICC robustness analysis

The poor inter-rater reliability of the global rating scale (ICC(2,1) = 0.146) reflects a well-documented limitation of subjective Likert-based clinical assessment instruments rather than a study-specific deficiency. Three supplementary analyses confirm that this does not threaten the study's conclusions.

##### S-Note 1a. Checklist and global rating measure partially distinct constructs

Pearson correlations between checklist and global rating scores were moderate at both time points (OSCE-1: $r = 0.32$, $p < 0.001$, $n = 444$; OSCE-2: $r = 0.54$, $p < 0.001$, $n = 366$), indicating that the two instruments capture partially overlapping but distinct aspects of clinical competence. The checklist assesses observable behaviours (whether specific actions were performed), while the global rating captures holistic impressions of communication quality. Per-examiner correlations ranged from $r = 0.42$ to $r = 0.75$, confirming that this moderate relationship holds across all raters (Supplementary Fig. S5a).

##### S-Note 1b. Low global ICC is a universal calibration problem, not a single-rater outlier

Leave-one-out analysis systematically removed each examiner from the dual-rated pairs and recalculated global rating ICC (Supplementary Table 9). Removing any single examiner did not improve ICC above 0.30 (range: 0.029–0.298), confirming that the low reliability reflects a universal anchoring problem inherent to the subjective scale rather than an individual outlier effect.

**Supplementary Table 9.** Leave-one-out ICC analysis for global rating (OSCE-1).

| Excluded examiner | ICC(2,1) | $n$ pairs | $\Delta$ from full |
|---|---|---|---|
| None (full model) | 0.146 | 146 | — |
| Zhang Haibin | 0.298 | 123 | +0.152 |
| Lu Xinyu | 0.113 | 77 | −0.033 |
| Ming Hao | 0.075 | 79 | −0.071 |
| Xu Danyang | 0.055 | 82 | −0.091 |
| Wang Lihong | 0.029 | 78 | −0.117 |

##### S-Note 1c. Sensitivity analysis: conclusions are identical regardless of scoring method

To verify that the low global rating ICC does not affect the study's primary conclusions, we repeated the between-group comparison using all three scoring methods as the outcome measure (Supplementary Fig. S5b). All three analyses yielded the same conclusion: no significant between-group difference in score change.

All three scoring methods yield non-significant between-group results (all $p > 0.26$), confirming that the study's conclusions are robust to the choice of outcome measure. The self-efficacy finding ($p = 0.034$) is entirely independent of OSCE rater reliability, providing an additional convergent outcome unaffected by this limitation.

**Supplementary Table 10.** Sensitivity analysis: between-group comparison by scoring method.

| Scoring method | ICC | Δ AI-SP | Δ Human SP | Δ Control | KW $p$ | $d_{AB}$ |
|---|---|---|---|---|---|---|
| Checklist (/14) | 0.717 | +1.4 | +0.9 | +1.0 | 0.483 | 0.38 |
| Global rating (/25) | 0.146 | +7.2 | +6.3 | +6.6 | 0.290 | 0.55 |
| Total (/39) | 0.400 | +8.6 | +7.3 | +7.6 | 0.266 | 0.52 |

### Supplementary Figure 5: Global rating ICC robustness

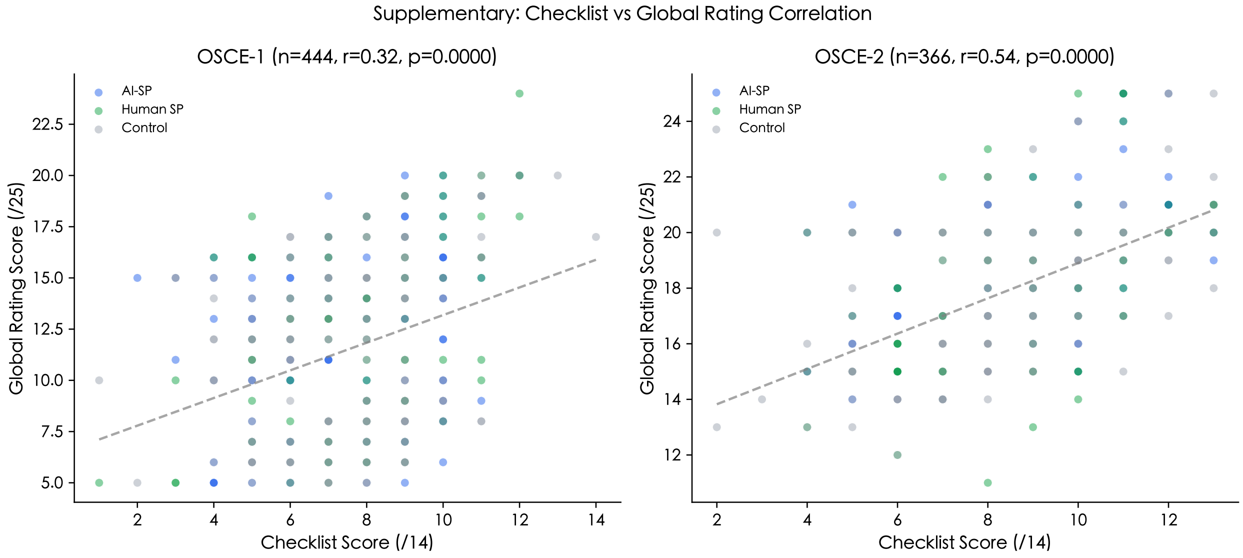


**Supplementary Figure 5a.** Checklist vs global rating score correlation at OSCE-1 ($r = 0.32$) and OSCE-2 ($r = 0.54$). The moderate correlation confirms that the two instruments measure partially distinct constructs; low global rating reliability does not compromise checklist-based conclusions.

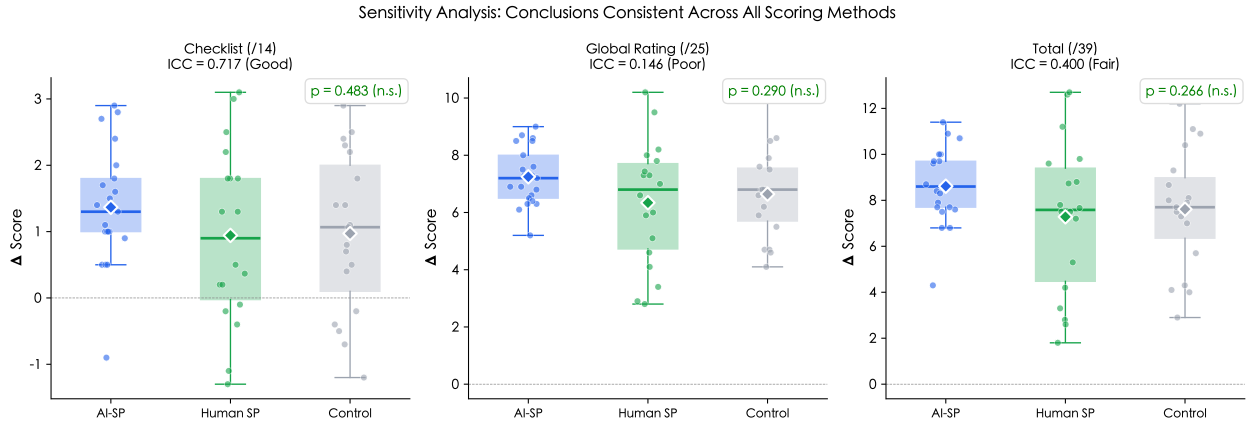


**Supplementary Figure 5b.** Sensitivity analysis: score change distributions by group across all three scoring methods. All methods yield the same non-significant conclusion (all $p > 0.26$), demonstrating that the study's findings are robust regardless of which scoring instrument is used.

### Supplementary Figure 6: Iceberg model of three-layer information architecture

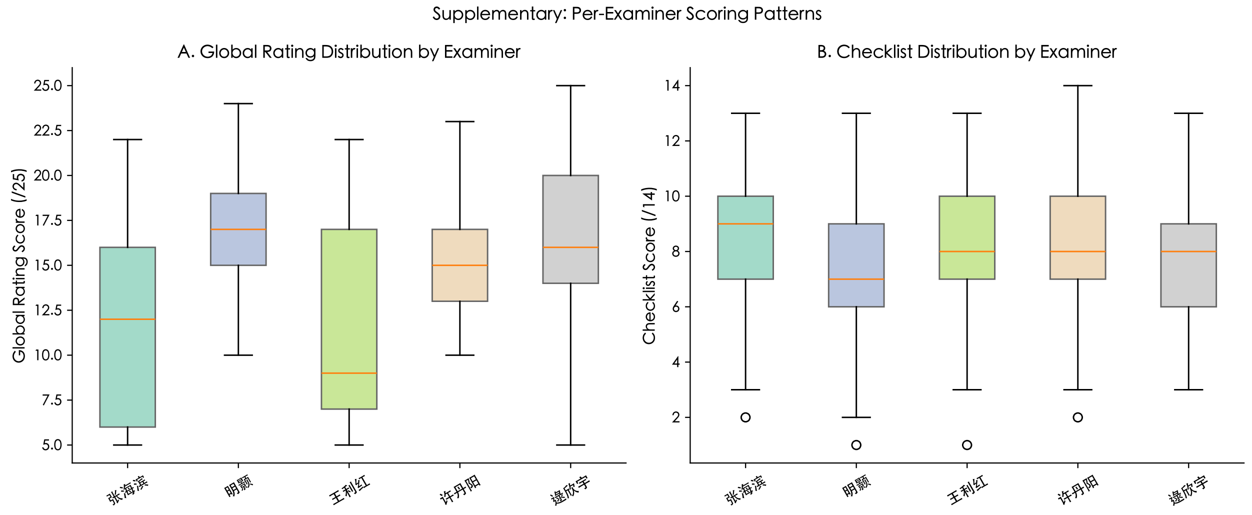


**Supplementary Figure 5c.** Per-examiner scoring distributions. Global rating distributions (left) show wide variation in examiner calibration (mean range: 6.7–16.0 in OSCE-1), while checklist distributions (right) are tightly clustered (mean range: 7.3–7.9), confirming that objective binary items resist subjective calibration drift.

#### Supplementary Table 2: Representative AI-SP system prompt structure

The following is the complete system prompt for Scenario 1 (laparoscopic cholecystectomy, 60-year-old male), presented in its original Chinese with annotated English translations of each structural section. All five scenario prompts follow this identical six-section template.

| Section | Content (translated summary) |
|---|---|
| **1. Patient identity** | Zhang Jianguo, male, 60, retired factory worker, high-school education. Personality: plain-spoken, somewhat stubborn, not talkative. Speech style: brief, direct, occasional dialectal expressions. |
| **2. Layer 1 items** (volunteered) | Three items disclosed spontaneously at consultation start: (a) gallstone surgery scheduled, recurrent pain; (b) hypertension, several years on medication; (c) no prior surgery. |
| **3. Layer 2 items** (disclosed on inquiry) | Eight items, each with explicit trigger and follow-up details. Examples: nifedipine SR 30 mg daily × 3 years (triggered by asking about specific medications); smoking half-pack/day × 30 years (triggered by lifestyle inquiry); sulfonamide allergy with urticaria (triggered by allergy history inquiry). Each item includes scripted "follow-up details" specifying dose, duration, and patient-appropriate qualifiers (e.g., "I don't know about liver enzymes"). |

| Section | Content (translated summary) |
|---|---|
| **4. Layer 3 items** (hidden, safety-critical) | Four items the patient does not recognise as surgically relevant: |
| | *(a) Self-purchased aspirin* — Takes aspirin 100 mg daily on neighbour's advice ("good for the heart"); considers it a supplement, not a medication. Triggered only when doctor explicitly asks about self-purchased or OTC medications.<br>*(b) Alcohol use* — 2–3 liang (100–150 mL) baijiu nightly × 30 years; minimised as "occasionally" unless specifically quantified.<br>*(c) Severe snoring/suspected OSA* — Spouse reports apnoeic episodes; patient dismisses as normal ageing. Triggered by sleep or snoring inquiry.<br>*(d) Loose molar* — Left lower molar mobile for 6 months; patient unaware of airway relevance. Triggered only by specific dental inquiry. |
| **5. Emotional model** | Baseline: tense but feigns nonchalance ("Surgery? Just get it done"). *If doctor is cold/hurried*: shorter answers, less forthcoming. *If doctor is patient*: gradually cooperative, willing to share more. *If lifestyle criticised*: defensive ("I've been fine all these years"). |
| **6. Behavioural rules & safety constraints** | (a) Colloquial Chinese matching a 60-year-old worker's register. (b) 1–2 sentences per response; no bracketed action descriptions. (c) Never volunteer Layer 2/3 information; disclose one hidden item per turn. (d) Provide scripted follow-up details when probed; say "I'm not sure" for unscripted details. (e) Counter-questions permitted ("What does this have to do with surgery?"). (f) **Safety block**: "You are a patient, not a doctor. Never provide medical advice, diagnoses, treatment plans, or drug recommendations. Never fabricate medical history beyond the script. Never break character." |

**Note.** The full Chinese-language prompts for all five scenarios are available in the data repository (`https://github.com/RegAItool/ai_sp_experiment`).

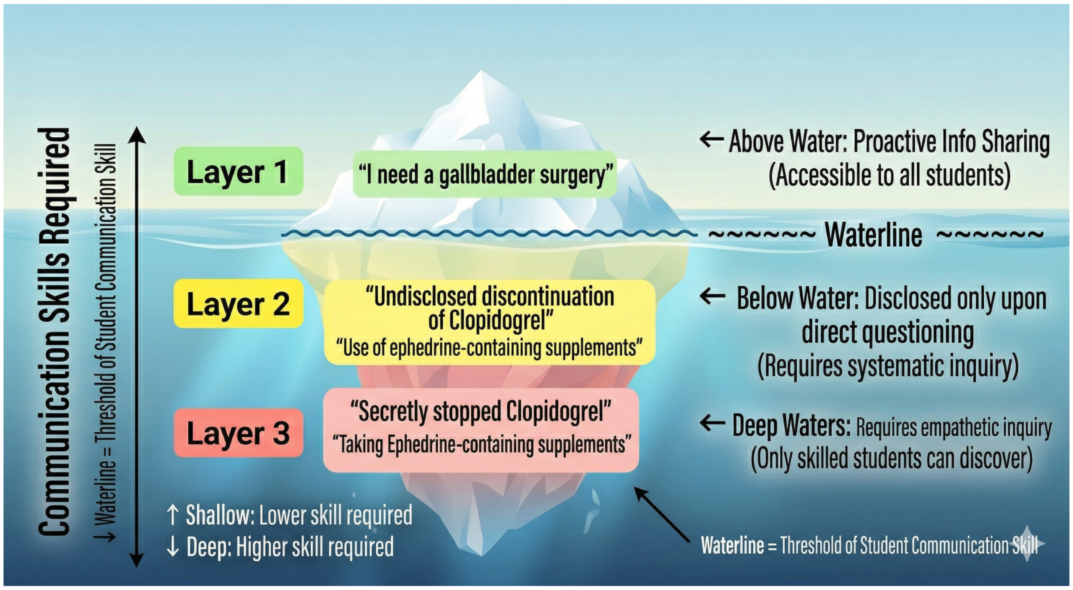


**Supplementary Figure 6.** The iceberg model of the three-layer information architecture. Layer 1 (surface) information is volunteered spontaneously by the AI-SP and accessible to all students. Layer 2 (prompted) information is disclosed only upon direct questioning, requiring systematic clinical inquiry. Layer 3 (hidden) information resides below the threshold of the patient's health literacy—the patient does not recognise its clinical relevance—and is disclosed only when the student demonstrates empathetic, targeted communication skills. The waterline represents the threshold of student communication competence: skilled interviewers access deeper information that remains hidden from novices. Example items shown are from Scenario 3 (knee replacement, 75F) and Scenario 4 (lumbar disc, 30M).
